# Active treatments outperform minimal intervention for adults with rotator cuff tendinopathy: a systematic review with predictive and network meta-analyses of complex interventions

**DOI:** 10.64898/2026.03.23.26349060

**Authors:** Rodrigo Rabello, Angie Fearon, Faiza Sharif, Bradley Stephen Neal, Phillip Newman, Simon Lack, Zubair Haleem, Victoria Tzortziou Brown, Kay Cooper, Paul Swinton, Dylan Morrissey

## Abstract

**OBJECTIVE:** To guide treatment of adults with rotator cuff tendinopathy (RoCuTe) by evaluating the relative efficacy of treatments, benchmarked against minimal intervention, for the co-primary outcomes of pain, function and quality-of-life (QoL) across short-, medium-, and long-term follow-up.

**DESIGN:** Systematic review with Bayesian predictive and network meta-analyses for synthesising complex interventions, guided by stakeholder involvement.

**FUNDING:** Private Physiotherapy Education Foundation (UK) Silver Jubilee Award.

**DATA SOURCES:** PubMed, Embase, Web of Science, CINAHL, and SPORTDiscus, searched to 22/8/2025.

**ELIGIBILITY CRITERIA FOR SELECTING STUDIES:** High-quality (PEDro score ≥7) randomised controlled trials comparing any intervention with another active or minimal intervention for patients clinically diagnosed with RoCuTe of either traumatic or insidious presentation; and reporting outcomes for pain, function, ± QoL.

**METHODS:** Title and abstract screening, full-text screening, and quality assessments were completed by two reviewers. Data extraction used the Elicit AI tool and was manually checked. Interventions were classified by treatment focus. Guided by patient and public involvement, pooled results from active interventions at short- (1 to ≤12 weeks), mid- (>12 weeks to <12 months) and long-term (≥12 months) were calculated for the primary analysis using Bayesian predictive meta-analysis models of within group change scores. Outcomes were benchmarked against an empirically derived minimal-intervention comparator (wait-and-see or sham). As a secondary analysis, network meta-analyses were conducted to synthesise relative effects and provide comparative rankings of active interventions. Risk of bias was assessed using the Cochrane Risk of Bias 2 tool, and certainty of evidence evaluated using GRADE.

**RESULTS:** We retained and analysed 140 high-quality studies that included 10,260 patients, 55.9% female, with a mean age of 48±8 years. Minimal interventions were associated with small short-term improvements, modest medium-term improvements and some regression in the long-term; in pain (0–100 scale: short=2.6; mid=23.3; long=21.1), function (standardised mean change (SMC): short=0.13; mid=0.87; long=0.76), and QoL (SMC: short=0.05; mid=0.33). At all timepoints, all active interventions with sufficient data were superior to minimal intervention for pain (0–100 scale: short=18.1–37.9 [14 categories]; mid=25.8–34.8 [8 categories]; long=30.8–45.0 [6 categories]), function (SMC: short=1.1–2.4 [14 categories]; mid=1.1–2.0 [11 categories]; long=1.0–1.8 [10 categories]), and QoL (short=0.8–1.7 [7 categories]; mid=0.9–1.8 [6 categories]). Certainty varied widely. Accordingly, three recommendation groups were defined based on the availability of comparative evidence and presence of higher-certainty findings. The strongest recommendation group included strengthening, range-of-motion exercises, complex interventions and movement pattern retraining.

**CONCLUSIONS:** A range of active treatments were superior to minimal intervention at each time point, so a wait-and-see approach should not be used, even in in the short-term. The most credible evidence was for interventions with a focus on strengthening, range-of-motion exercises, movement pattern retraining, and complex interventions. Clinicians should prioritise active management and deploy personalised clinical reasoning to tailor treatment to patient preferences and the available resources.

**SYSTEMATIC REVIEW REGISTRATION:** *PROSPERO CRD42024584126:* WHAT IS ALREADY KNOWN ON THIS TOPIC

- Rotator Cuff Tendinopathy is a common and troublesome condition, leading to pain, decreased function and quality-of-life that can be severe and is not self-limiting.
- Many treatment options have proven efficacy, but the optimal management is unclear, leading to treatment variability and unsatisfactory outcomes. WHAT THIS STUDY ADDS

- We estimated the improvements in pain, function and quality-of-life resulting from minimal interventions (wait-and-see or sham), confirming that the condition is not self-limiting. Several active treatments had sufficient data showing they are superior to minimal intervention in the short (3 months), mid (6 months) and long-term (12 months) Although the network meta-analysis did not provide a clear hierarchy between active treatments, certainty of evidence and data volume suggest that strengthening, range of motion exercises, movement pattern retraining and complex interventions are the treatments of choice.

## INTRODUCTION

Shoulder problems account for 2.4% of all consultations with general practitioners in the UK,^1,2^ with 1% of adults presenting with new shoulder pain every year.^3,4^ Among these, approximately 70% are related to the rotator cuff^5^ and are labelled using similar terms, including rotator cuff tendinopathy, rotator cuff related shoulder pain, shoulder impingement syndrome, subacromial pain syndrome,^6^ Hereafter, we refer to these conditions collectively as rotator cuff tendinopathy (RoCuTe). There are currently no definitive treatment recommendations for RoCuTe, and persistent symptoms are commonly reported even after 5 years,^7^ contributing to a substantial health burden and associated direct and indirect costs.^8^

Although many randomised controlled trials (RCTs) have investigated the effects of different RoCuTe treatment options, there is no consensus on which approach is best. Previous systematic reviews that have synthesised the available evidence are limited by focusing on the efficacy of single treatments or only a narrow subset of interventions.^9–11^ Further, while interventions in RCTs are usually labelled according to a primary treatment, these are often delivered as complex interventions composed of multiple treatments (i.e., components of the intervention). This complexity makes pooling studies challenging and can require discarding informative data, as highlighted in a recent network meta-analysis.^12^ Moreover, findings from previous systematic reviews are often at high risk of bias or misleading interpretation due to the inclusion of low-quality studies.

Taken together, these limitations mean that current evidence does not yet provide clinicians with clear, high-quality guidance across the full range of conservative options for RoCuTe. A synthesis of RCT evidence that benchmarks active interventions against a credible, data-driven minimal-intervention outcome estimate is therefore needed to inform clinical recommendations across these conservative treatment options. Given that interventions are often complex and variably composed, analyses must accommodate multiple components and limited direct comparisons. Accordingly, we applied a predictive meta-analytic framework to estimate the absolute benefit of active treatments over the minimal-intervention benchmark in future studies, and a network meta-analysis to integrate direct with indirect evidence to enable comparative evaluation across interventions. Guided by stakeholder engagement, we aimed to identify: (1) which treatments for adults with RoCuTe are superior to minimal intervention; and (2) whether a treatment hierarchy can be inferred, based on high-quality RCTs at low risk of bias, for the co-primary outcomes of pain, function, and quality-of-life (QoL), across short-, medium-, and long-term follow-up.

## METHODS

### Overview and deviations from protocol

This systematic review was conducted following the Cochrane recommendations and is reported according to the Preferred Reporting Items for Systematic reviews and Meta-Analyses (PRISMA) guidelines and the network meta-analysis, complex intervention, and search reporting extensions (Supplementary S1 and S2).^13–15^ Operational definitions of terms used are presented in Supplementary S3. This review was pre-registered with PROSPERO (CRD42024584126), and four protocol deviations were implemented due to the volume and characteristics of the available data. We did not analyse cost outcomes or binary outcomes, as insufficient relevant data were found. Rather than dual independent data extraction, the Elicit tool (Elicit Research PBC, Covina, USA) was used for primary extraction, which was then manually checked and completed by one researcher. For initial quality analysis, we added an additional item to the PEDro score to account for conflicts-of-interest. We also conducted sensitivity analysis focusing on interventions with a single intervention and for those with no adjuncts, as opposed to the original planned sensitivity analyses for patient subgroups or for higher-quality studies.

### Patient and public involvement (PPI)

Prior to undertaking this review, we conducted two PPI sessions with three patients with RoCuTe (two females, one male) and six clinicians (three physiotherapists and three surgeons), as well as structured meetings within our interdisciplinary research team. These activities informed the selection of pain, function, and QoL as the primary outcomes, refined eligibility criteria, and shaped the systematic review research questions, including the comparison to minimal intervention as the primary benchmark. Additionally, one patient was included as part of the authorship group to elevate the patients’ voices across the completion of the review.

### Search strategy and information sources

PubMed, Embase, Web of Science, CINAHL, and SPORTDiscus were searched (the latter two through EBSCO), without language restrictions or filters, from inception until August 22^nd^, 2025 (initial search conducted on July 15^th^, 2024), for eligible RCTs. Search strategy was created based on terms used in previous reviews and reviewed by a group of experienced systematic reviewers. MeSH and selected terms for study design and population were searched for in all fields (Table 1). Other PICO elements were not restricted, as there were no limits to possible interventions, comparators, or outcome terminology. Subsequently, one investigator searched for additional records by conducting forward and backward citation tracking of included studies through the Scopus database and by screening publications from known researchers in the field. No additional methods were used to identify records.

**Table 1.** Search terms used for identifying records

|  |  |
| --- | --- |
| Study design terms | randomised controlled trial OR Randomized Controlled Trial<br>OR random allocation OR Controlled Clinical Trial* OR Clinical<br>Trial* OR controlled trial* |
| Population - RoCuTe | Rotator cuff related shoulder pain OR Shoulder impingement<br>syndrome OR Subacromial impingement syndrome OR<br>Rotator cuff tend* OR Shoulder burs* OR Subacromial burs*<br>OR Subacromial pain syndrome OR Rotator cuff disease OR<br>Rotator cuff tear OR Shoulder tend* OR Supraspinatus tend*<br>OR Infraspinatus tend* OR Teres minor tend* OR<br>Subscapularis tend* OR Painful arc |
| Full search for each database and the breakdown of records identified through each term<br>are presented in Supplementary S4 and S5. |  |

### Eligibility criteria and selection process

Records were exported into COVIDENCE (Veritas Health Innovation, Melbourne, Australia), where duplicates and records automatically classified as not being RCTs were removed, a process shown to identify over 99.5% of references correctly.^16^ Two independent reviewers (RR, and FS or DM) reviewed all titles and abstracts according to the inclusion criteria (Table 2), with conflicts resolved by discussion and consensus. Relevant full texts were evaluated by a single reviewer (RR) against the inclusion criteria, with a second reviewer (FS) screening 20% of full-texts and confirming all exclusions of the remaining 80%, with a third reviewer (DM) available to resolve conflicts.

**Table 2.** Eligibility criteria.

| Inclusion | Exclusion |
| --- | --- |
| • Peer-reviewed randomised controlled trial<br>• Comparison of at least one conservative treatment for RoCuTe with any other treatment or minimal intervention (wait and see, placebo or sham)<br>• Adults aged 18 and over<br>• Primary diagnosis of: Subacromial impingement syndrome, rotator cuff tendinopathy, subacromial pain syndrome, rotator cuff tendinitis, rotator cuff related shoulder pain.<br>◦ Secondary diagnosis of small partial and full thickness rotator cuff tears, calcific tendinitis, subacromial bursitis, non-radiating neck pain were allowed<br>• Diagnosis made by a clinician using recognised clinical methods including examination but not requiring imaging confirmation<br>• Traumatic or insidious presentation at any stage.<br>• Contain at least one outcome related to QoL, pain or function measured for a minimum of one week | • Non-randomised trials, non-peer reviewed reports, feasibility or pilot studies.<br>• Comparison of two or more surgical techniques, with no conservative or wait and see arms<br>• Studies including children and adolescents<br>• Follow-up shorter than one week<br>• Different primary diagnosis or where the data for participants with RoCuTe cannot be clearly extracted in isolation: such as osteoarthritis, fractures, frozen shoulder, medium or massive rotator cuff tears, periarthritis, diabetes, spinal cord injury, radiating neck pain<br>• Participants that had received shoulder surgery prior to inclusion in the study<br>• Diagnosis only through imaging or self-reporting<br>• Shoulder pain because of cerebrovascular accident or systemic inflammatory conditions.<br>• Trials where the sole outcome is radiological or other mechanistic outcomes (e.g., acromio-humeral distance, EMG, scapular position)<br>• Reports with data, already included in the review through a different report<br>• Doubts regarding ethical approach to publication (i.e., statements of concern or multiple publications of same data) |

### Study quality and risk of bias

In accordance with previous methods,^17,18^ reports that met the inclusion criteria were evaluated using the PEDro scale.^19,20^ One additional risk of bias criterion was added to account for conflicts of interest, such as injection studies funded by a pharmaceutical company, or where no conflict-of-interest statement was reported. Only reports that scored ≥8/11 points were considered to have adequate methodological quality. Additionally, studies scoring less than 6 on the six items common to the PEDro scale and the Cochrane Risk of Bias Tool, plus the conflict-of-interest item, were excluded. All reports were scored by one reviewer (RR); those at the inclusion threshold (i.e., total PEDro score of 7 and 5 points on the shared items) were evaluated by a second reviewer (FS or DM), with conflicts resolved by discussion.

### Data collection

A detailed data extraction codebook (Supplementary S6) was constructed, and manual extraction on 30 test reports was undertaken. Data extraction was then completed using the Elicit tool (Elicit Research PBC, Covina, USA), where each manuscript was uploaded and prompts were provided for the extraction of relevant data. All data extracted by the tool were manually confirmed by one investigator (RR) and supplemented where required. Published study protocols, supplementary material, and study registrations were also manually checked to complete or correct extracted data. When data were unavailable or unclear within a manuscript, authors were contacted by email up to two times. When results were reported using both intention-to-treat and per-protocol analyses, the latter was extracted. Values from studies reported only in figures were estimated by digitisation using ImageJ (National Institute of Health, Bethesda, MD, USA).

Data extracted included participant demographics (e.g., age, symptom duration, symptom severity, and BMI), study methods (e.g., outcomes, time points), detailed intervention characteristics (elements, duration, frequency, adherence, fidelity, and development), and group mean and distribution values for all outcomes of interest (Supplementary material 4)

### Outcomes

Guided by PPI meetings, only outcomes related to pain, function, and QoL were extracted, with a maximum of two outcomes per category for a given report (primary outcome plus one additional outcome, if applicable). A pre-determined hierarchy was applied to select outcomes where no hierarchy was reported in the report: Pain and Disability Index (SPADI), Disabilities of the Arm, Shoulder and Hand (DASH) questionnaire, Constant-Murley Shoulder Outcome Score (CM), Oxford Shoulder Score (OSS) or other available function tools; EQ-5D, Short Form Health Survey (SF-36), Western Ontario Rotator Cuff (WORC) Index, or other available QoL tools. When multiple domains of a given outcome were reported, we selected the overall result when combining domains was recommended by the developers (e.g., SPADI and CM). In cases involving QoL outcomes, where domains were not meant to be combined, (e.g., SF-36 and Nottingham Health Profile) we extracted each domain as a separate outcome.

### Interventions

When interventions contained multiple treatments (e.g., injection plus stretching), information regarding treatment type, duration, frequency, intervention development, adherence, and fidelity was extracted for each treatment separately, with intervention foci (up to a maximum of four) recorded for each study arm. Exercise type, setting (home or supervised), number of sets, number of repetitions, and intensity were further extracted for strengthening elements. Strengthening elements were also categorised as poorly or well-reported, and as having sufficient or insufficient load (Supplementary S7). Education was distinguished from information provision as structured, well planned, and having indications of knowledge assessment.

### Timepoints

Follow-up data were extracted for the short-term (≥1 week and ≤12 weeks), medium-term (>12 weeks and <12 months) and long-term (≥12 months) following the start of treatment. When multiple timepoints were provided within one period, data from the longest available timepoint within that period were extracted (e.g. 12 weeks over 8 weeks), with the exception of long-term follow-up, where the 12 months timepoint was preferred over longer follow-up durations.

### Treatment classification

Study arms were classified as minimal or active interventions, which were further categorised according to treatment foci. Complex interventions were also identified.

#### Minimal intervention

Minimal intervention was chosen as a pragmatic benchmark rather than a pure estimate of untreated natural history, recognising that even sham or minimal-contact interventions can induce improvement through contextual and expectancy effects. To obtain an estimate of the natural history, treatment arms that contained a minimal intervention were collated. These were categorized as: (1) wait-and-see (approach where the participants do not receive a systematic intervention); (2) inert sham (sham version of the experimental group, involving only a light touch to the patient and not likely to have a physiological effect); (3) sham (sham version of the experimental group, involving more intense treatment likely to have physiological effect); (4) sham combined with active secondary intervention (sham treatment combined with active treatments as an adjunct). For this benchmarking purpose only, data from the relevant arms of low-quality studies were also included when they contained wait-and-see, inert sham or sham comparators, to maximise estimation accuracy and precision in the presence of sparse data.

#### Active intervention classification

The treatment focus of active interventions was determined based on the study title, aim, and group names. Each intervention could be assigned up to four treatment foci, with other treatments classified as adjuncts. A detailed description of the procedures used to determine treatment foci and adjuncts is provided in Supplementary S8.

#### Complex interventions

Interventions with three or four treatment foci of treatments judged a priori to be plausible based on existing evidence (including being superior to minimal intervention) and clinical rationale were analysed according to their individual foci and additionally classified as “complex interventions” (Supplementary S9).

#### Highly complicated multi-treatment interventions

Interventions with five or more treatment components (plausible or not) were considered too complex to permit attribution of observed effects to individual components. For this classification, information provision or anaesthetic administered in combination with injection were not counted as separate treatment components (Supplementary S9).

#### Pooling of studies

Interventions were pooled according to their focus at the highest level of resolution feasible, defined by having at least two studies with a minimum of 100 participants (Supplementary S10). For the purposes of the meta-analysis, we combined treatments where possible according to their clinical interest, plausibility, and underlying rationale, and conducted sensitivity analyses at higher levels of aggregation when appropriate. For example, motor control, proprioception and stability exercises were combined into the category “Movement Pattern Retraining” and dry needling and acupuncture were combined into “Needling”. Interventions were assigned to multiple treatment categories if they had multiple foci. Treatments that could not be combined were entered into the meta-analysis individually, whereas surgical treatments were not entered as we excluded any surgical vs surgical studies, resulting in their underrepresentation in the present review. Thus, surgical arms, as well as trials containing only broad descriptions of exercise or physiotherapy were classified as “other” and not included in the meta-analyses.

### Data synthesis

#### Data management

For pain outcomes, absolute mean change scores were calculated on a 0–100 scale, with negative values indicating improvement. For function and QoL, outcomes were extracted on their original scales and converted to standardised mean change scores (SMCs) to enable pooling across outcome measures. All values triggering verification (defined as outside of 0.3 to 2.0 SMC or outside of −40 to −10 change in pain) were manually confirmed by rechecking the original reported results.

#### Minimal-intervention analysis

We generated Bayesian predictive outcome distributions for the four levels of minimal intervention—1) wait-and-see; 2) inert sham; 3) sham; or 4) sham with active secondary intervention—to represent the effects expected in future studies using these interventions. These predicted distributions were compared across categories, and their relative ordering was summarised using the Surface Under the Cumulative Ranking curve (SUCRA). This produced a clear hierarchy across the four minimal-intervention categories, from wait-and-see through to sham with a plausible secondary intervention (Figure 2 and Supplementary S11). Therefore, we determined the minimal intervention benchmark for comparison with active treatments by starting from the first level (wait-and-see) and progressively adding studies from subsequent levels until at least two studies were available for each outcome-timepoint combination.

#### Minimal vs active intervention comparison

Predicted outcome distributions expected in future studies were obtained using Bayesian random-effects meta-analyses, employing a Student-t likelihood to allow for heavy-tailed error distributions. Posterior predictive simulations were then used to generate the expected distribution of outcomes in a new trial, and the proportion of simulated outcomes (m=10,000 iterations) that exceeded the minimal-intervention benchmark was taken as the probability that a specific active treatment would be superior. The best estimate of treatment efficacy was quantified directly from the posterior distribution of the pooled mean from the meta-analysis. The probability that a future study including the active treatment would exceed the minimal clinically important change (MCIC) was calculated using the posterior predictive simulations. MCIC thresholds were defined a priori as −15 units for pain,^21^ +0.6 SMC for function^21–23^ and +0.4 SMC for QoL.^22^

Three levels of analyses were conducted based on the intervention focus. In the main analysis, all study arms with that focus, whether delivered alone or in combination with other treatments, were assigned to the treatment category. A secondary analysis (sensitivity 1) included study arms with a single focus. A third analysis (sensitivity 2) included only study arms with that single focus and no adjunct treatments. Results from these sensitivity analyses are presented in the electronic Supplementary File.

Further sensitivity analyses were conducted to evaluate higher-resolution treatment categories when sufficient data were available: high-intensity strengthening, low-intensity strengthening, well-developed complex interventions, poorly-developed complex interventions, and non-steroid injections (PRP, hyaluronic acid, botulinum toxin, glucose, anaesthetic, ozone, and tenoxicam).

#### Outlier handling

We screened individual change values for each outcome and time point using a robust, median-based z-score (MAD-based). Values were flagged for verification and, where appropriate, removed when the robust z-score exceeded a conservative absolute threshold (3.5), following conventions described by Iglewicz & Hoaglin.^24^

#### Network meta-analyses

To enable comparison across treatment options, we conducted Bayesian network meta-analyses that integrated both direct head-to-head evidence and indirect comparisons arising from the network structure. Networks were specified using arm-based parameterisation, with strengthening interventions used as the reference comparator because they were the most frequently evaluated treatment. Only control arms (i.e., minimal intervention), single-focus active interventions, and eligible complex interventions (entered as a single node) were included to preserve interpretability of treatment effects.

The principal summary measure was the difference in mean change from baseline (pain; 0–100 scale) and standardised mean change (function and QoL), summarised as posterior median treatment effects with 95% credible intervals. In addition to pairwise relative effects, treatments were summarised using posterior ranking probabilities and SUCRA values.

Multi-arm trials were modelled within a single likelihood to account for within-study correlation. Between-study heterogeneity was modelled using a random-effects structure with a shared heterogeneity parameter (τ) within each outcome–timepoint network. Regularising priors were applied to stabilise estimation in sparse networks. For pain (0–100 scale), class effects were assigned Normal(0, 7) and heterogeneity τ ∼ half-Normal(0, 2.5), with study baselines θ ∼ Normal(0, 20). For function and QoL (standardised mean change), class effects were assigned Normal(0, 0.5), heterogeneity τ ∼ half-Normal(0, 0.5), and study baselines θ ∼ Normal(0, 1.5). Arm-level random deviations were modelled via δ_raw_ ∼ Normal(0, 1) and scaled by τ. Model fit was assessed using the posterior deviance statistic, calculated as the sum of squared standardised residuals across contributing study arms, and interpreted relative to the number of study arms. Inconsistency was evaluated by comparing a consistency model (u = 0) to an inconsistency model that added a global inconsistency deviation for each non-baseline arm, u_a_ = κ·u_raw,a_, with κ ∼ half-Normal(0, 0.5) and u_raw_ ∼ Normal(0, 1), and u anchored to zero for each study baseline arm. Evidence for inconsistency was judged by (i) reductions in posterior deviance under the inconsistency model relative to the consistency model and (ii) the posterior distribution of κ, where larger values indicate greater disagreement between direct and indirect evidence.

All analyses were conducted in R (version 4.5.2). Bayesian models were implemented in Stan and/or via brms interfacing with Stan for Markov chain Monte Carlo sampling. Uncertainty was quantified using appropriate credible intervals (75 or 95%).

### Certainty of Evidence (GRADE)

Certainty of evidence was assessed in accordance with the GRADE framework and evaluated at the level of our primary analysis (comparison of active interventions with the benchmarked minimal-intervention estimate) and secondary analysis (network meta-analysis of relative treatment effects). Certainty was initially rated as high and was downgraded according to predefined criteria.

Risk of bias was evaluated using the Cochrane Risk of Bias 2 (RoB2) tool, with excerpts extracted via the Elicit tool to guide application of the signalling questions and elaboration criteria. One investigator (RR) completed the RoB2 assessments for all reports, and final judgements were reached by consensus with two additional investigators (DM and AF) in cases of uncertainty. Across the body of evidence, reports were classified by study and domain as having low risk of bias, some concerns, or high risk of bias. Certainty was downgraded one level if ≥50% and <75% of pooled data came from studies with high risk of bias or some concerns, and downgraded two levels if ≥75% of pooled data came from studies with high risk of bias or some concerns.

Publication bias was assessed by sampling trial registrations from the International Clinical Trials Registry Platform across three representative periods (2009–10, 2015, and 2020). For each period, a random sample of 15 eligible trials was screened for intervention focus and publication status (Supplementary S12). Publication bias was suspected when unpublished trials outnumbered published trials for a given treatment category, and this resulted in downgrading certainty of evidence by one level.

For the predictive meta-analyses, imprecision was judged based on the magnitude of the posterior probability that an active intervention would be superior (or inferior) to the minimal-intervention benchmark. For superiority, probabilities of 0.50–0.60 were interpreted as very low certainty, 0.60–0.75 as low certainty, 0.75–0.90 as moderate certainty, and ≥0.90 as high certainty of superiority. Probabilities below 0.50 were interpreted symmetrically as evidence of inferiority. Probabilities classified as low or very low certainty were considered to reflect serious imprecision and resulted in downgrading by one level. Inconsistency was assessed using the posterior median between-study standard deviation. Values exceeding 7.5 points on the 0–100 pain scale or 0.4 SMC for function and QoL were considered indicative of serious inconsistency and downgraded one level.

For the network meta-analyses, imprecision was judged based on the magnitude and uncertainty of relative treatment effects compared with strengthening, alongside the stability of posterior ranking estimates. Downgrading was informed by whether 95% credible intervals crossed no difference, the extent to which effects were small relative to typical within-arm changes, and the dispersion of posterior ranks and SUCRA values. Greater imprecision and instability in relative effects and rankings resulted in more severe downgrading. Inconsistency was assessed based on the magnitude of between-study heterogeneity and agreement between direct and indirect evidence. Judgements were informed by the posterior distribution of the heterogeneity parameter (τ) and by comparing model fit between consistency and inconsistency models using residual deviance. Downgrading was applied when heterogeneity was large or when allowing for inconsistency meaningfully improved model fit, indicating disagreement between direct and indirect evidence.

### Recommendations

Based on strength of evidence including the number of comparisons available across outcomes and timepoints, and the GRADE certainty of evidence, we divided our recommendations into three groups plus education, typically deployed alongside all other interventions ; 1) Complete evidence with higher certainty – Data available for most outcome-timepoint combinations, with at least two comparisons demonstrating high certainty of high or moderate superiority; 2) Incomplete evidence or no higher-certainty comparisons – Insufficient data for ≥3 outcome-timepoint combinations and/or no comparisons demonstrating high or moderate superiority with high certainty of evidence; and 3) Very limited evidence – Insufficient data for most outcome-timepoint combinations.

## RESULTS

### Study and population characteristics

Details of the screening process are shown in the PRISMA diagram (Figure 1). In total, 8,323 records were identified and screened, of which 307 met the inclusion criteria, with 140 judged to be high-quality and included in the review (list of included studies for low-quality and other reasons are in Supplementary S13 and S14). An additional 10 low-quality studies that included a minimal-intervention arm were incorporated for benchmarking purposes. For the pain outcome, 109 studies contributed data to at least one treatment category and were entered into the meta-analyses for at least one timepoint. For function, 116 studies were included, and 27 studies contributed data for QoL outcomes. Across all included studies, 10,382 participants (55.9% females) were included, with a mean age of 48 ± 8 years, symptom duration of 15.4 ± 13.1 months, body mass index of 26.1 ± 2.1 kg/m², and symptom severity of 50.5 ± 14.6 on a normalised scale derived from SPADI, DASH, Constant-Murley score, and other available function or pain outcomes.

**Figure 1.**
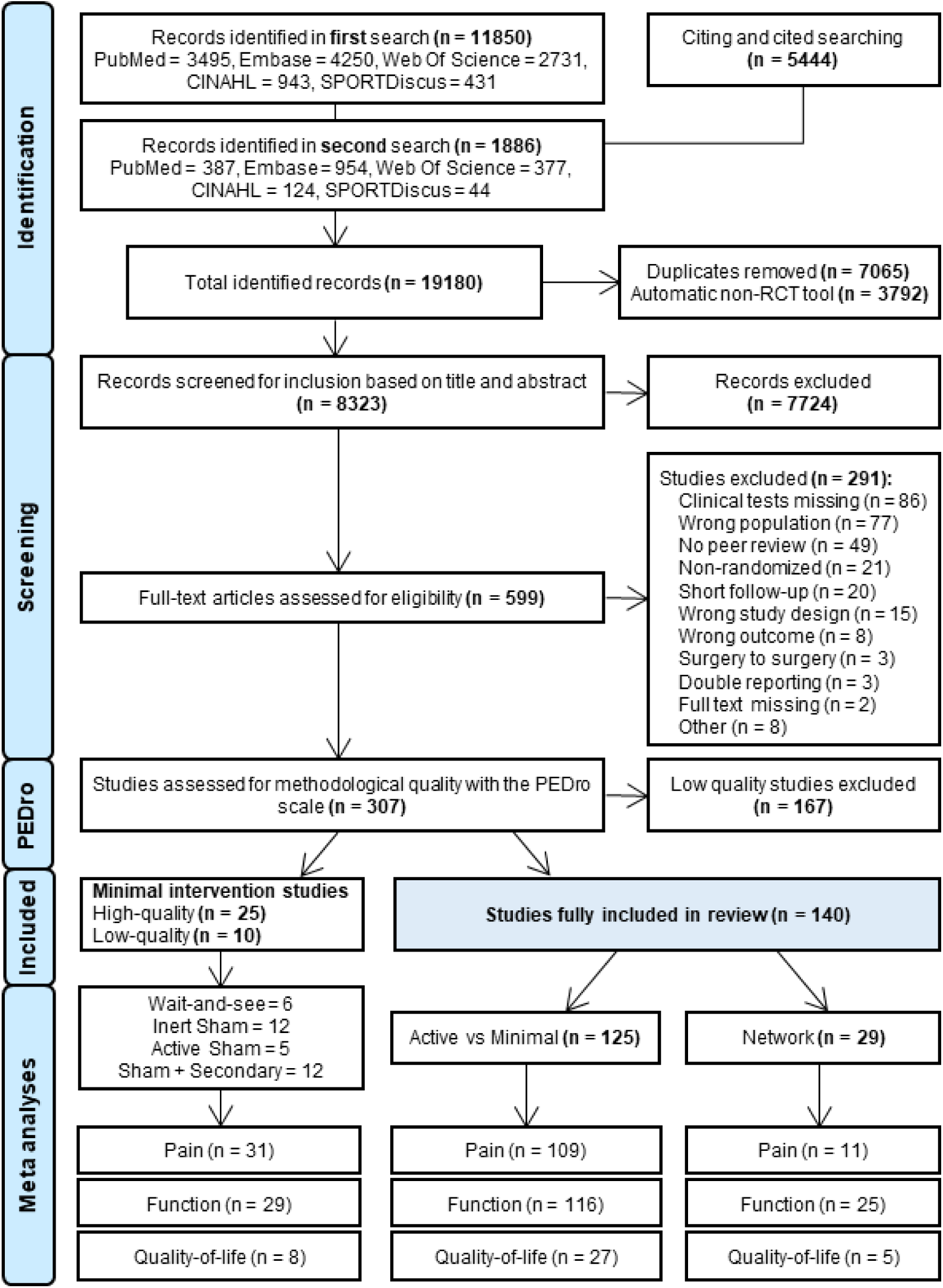
– PRISMA flow diagram for screening and selection

## GRADE

No serious concerns regarding indirectness were identified, as the included populations, interventions, and outcomes were directly relevant to the review question. Certainty of evidence was not downgraded for risk of bias, as the contribution of studies at high risk of bias did not exceed prespecified thresholds. Publication bias was suspected of injection therapies (including steroid and other injections) and acupuncture/dry needling. No publication bias was identified for other treatment categories.

For the predictive meta-analyses, inconsistency and imprecision were assessed separately for each outcome and timepoint. Serious inconsistency, defined by large between-study heterogeneity, was identified in a minority of treatment categories for pain (short: 2/14; mid: 1/8; long: 0/6), but was more common for function (short: 4/14; mid: 2/11; long: 7/10) and QoL (short: 3/7; mid: 4/6). Where present, serious inconsistency resulted in downgrading of the certainty of evidence by one level. Serious imprecision, based on low or very low posterior certainty of superiority, was uncommon for pain (short: 0/14; mid: 0/8; long: 2/6) and function (short: 1/14; mid: 0/11; long: 0/10), but was more frequent for QoL (short: 2/7; mid: 3/6). Evidence judged to have serious imprecision was downgraded by one level.

For the network meta-analyses, very serious imprecision was identified across all outcomes and timepoints assessed, reflecting wide credible intervals and unstable ranking estimates. No serious inconsistency was identified for pain at short-term follow-up or for function at short- and mid-term follow-up. Some concerns regarding inconsistency were identified for short-term function and short-term QoL, whereas no serious inconsistency was identified for other comparisons.

### Minimal-intervention analyses and benchmarking

Thirty-five studies included minimal-intervention arms, including 10 classified as low-quality. These comprised 6 arms of wait-and-see, 12 of inert sham, 5 of sham, and 12 of sham combined with active secondary intervention (Supplementary S15). Improvements from baseline increased consistently with the ranking of minimal-intervention levels (Figure 2).

**Figure 2.**
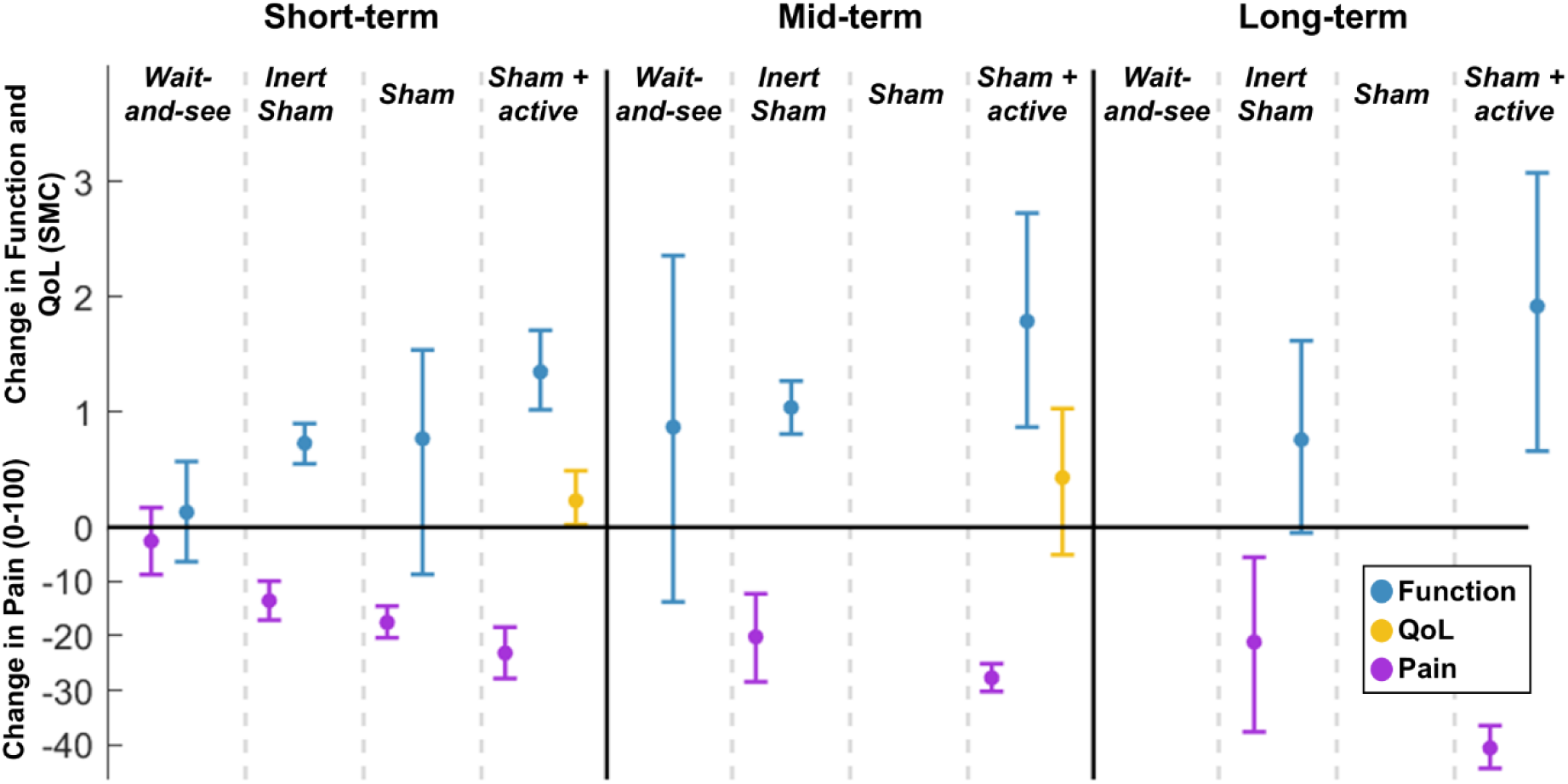
Change from baseline across the four levels of minimal intervention (bottom panel: pain; top panel: function and quality-of-life). Intervals represent 75% credible intervals and summarise the central mass of the posterior distributions. Although displayed as line segments, these intervals reflect underlying distributional information, which is illustrated in full in the Supplementary S11. QoL: Quality-of-life; SMC: Standardised mean change.

To establish the minimal-intervention benchmark, we used wait-and-see alone for short-term pain and function, and mid-term function; wait-and-see combined with inert sham for mid-term pain and short-term QoL; inert sham alone for long-term pain and function; and inert sham combined with sham plus plausible secondary intervention for mid-term QoL. No sufficient minimal-intervention data were available for long-term QoL. The benchmarked minimal interventions were associated with small short-term improvements, larger mid-term improvements, and partial regression at long-term follow-up, particularly for pain and function (Figure 2). Improvements increased consistently across the hierarchy of minimal-intervention categories, supporting their use as a graded benchmark for comparison with active interventions (Supplementary S11).

### Comparison between active and minimal interventions

Comparison to the minimal-intervention benchmark was made for interventions designated as complex and for 13 active treatments identified as a treatment focus (detailed analyses with summaries and domain GRADE analyses presented in Supplementary S16). Data were included from 121, 44, and 19 studies in the short-, mid-, and long-term follow-up, respectively (Active intervention characteristics presented in Supplementary S17). Across the 76 total comparisons (pain = 28, function = 35, QoL = 13), the median posterior probability of superiority exceeded 0.5 in all cases (mean ± sd = x ± y), indicating consistent superiority relative to the minimal-intervention benchmark (Figure 3).

**Figure 3.**
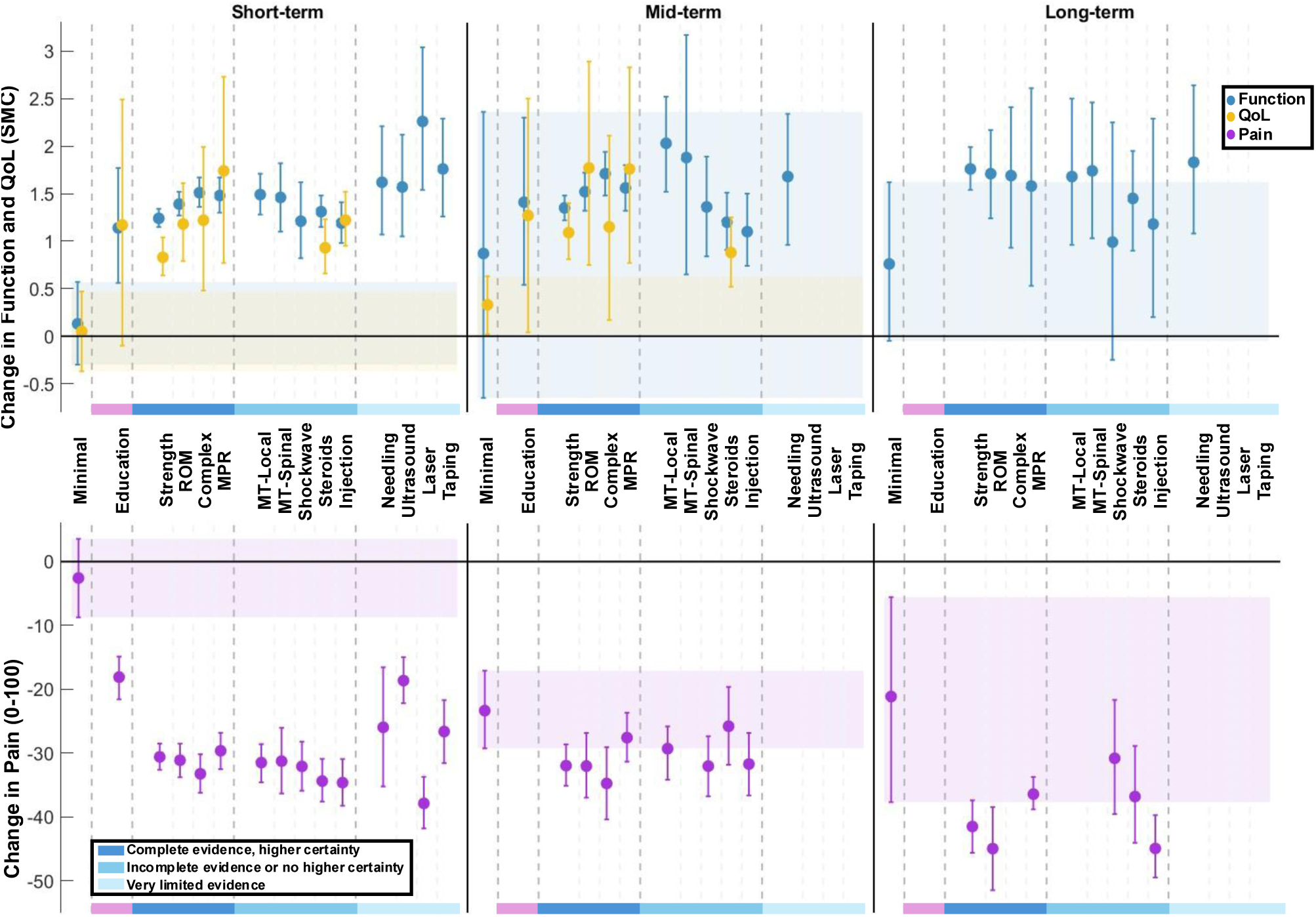
Change from baseline across active intervention focuses and minimal-intervention benchmarks (top panel: pain; bottom panel: function and quality-of-life). Recommendation groupings are separated by grey vertical lines and coloured bars on x-axis. These groups denote recommendation level based on evidence availability and higher-certainty comparisons based on GRADE assessment. Intervals represent 75% credible intervals and summarise the central mass of the posterior distributions. Although displayed as line segments, these intervals reflect underlying distributional information, which is illustrated in full in the Supplementary Material. QoL: Quality-of-life; SMC: Standardised mean change. Minimal intervention benchmarks comprise wait-and-see only for short-term pain and function and mid-term function; wait-and-see plus inert sham for mid-term pain and short-term quality-of-life; inert sham only for long-term pain and function; and inert sham plus sham combined with active secondary intervention for mid-term quality-of-life. Sufficient minimal intervention data were not available for long-term quality-of-life.

In the short-term, point estimates from the pooled mean change scores indicated improvements ranging from 18.1 to 37.9 on the 0-100 pain scale, 1.1 to 2.3 SMC for function, and 0.83 to 1.7 SMC for QoL. Of the 35 comparisons, certainty of superiority was judged as high for 10 comparisons (pain: 10 (71%), none for function or QoL), moderate for 22 comparisons (pain: 4 (29%); function: 13 (93%); QoL: 5 (71%)), and low for 3 comparisons (function: 1 (7%); QoL: 2 (30%), none for pain); no comparisons were rated as very low certainty.

In the mid-term, point estimates indicated improvements ranging from 25.8 to 34.8 on the 0-100 pain scale, 1.1 to 2.0 SMC for function, and 0.9 to 1.8 SMC for QoL. Of the 25 comparisons, none were rated as high certainty, 3 comparisons were rated as moderate (QoL: 3 (50%), none for pain or function), 14 as low certainty (pain: 6 (75%); function: 5 (46%), QoL: 3 (50%)), and 8 as very low certainty (pain: 2 (25%); function: (55%), none for QoL).

In the long-term, point estimates indicated improvements ranging from 30.8 to 45.0 on the 0-100 pain scale, and 1.0 to 1.8 SMC for function. As insufficient data were available to establish a long-term QoL minimal-intervention benchmark, QoL comparisons at this timepoint were not conducted. Of the 16 comparisons, none were rated as high certainty; 2 were rated as moderate (pain: 2 (33%); none for function), 12 as low certainty (pain: 4 (67%); function: 8 (80%)), and 2 as very low certainty (2 function: 2 (20%), none for pain).

The recommendation group that had complete evidence with higher certainty included strengthening, range-of-motion exercises, complex interventions, and movement-pattern retraining. These treatment categories provided sufficient data for most outcome-timepoint combinations and showed superiority (high or moderate) with high certainty of evidence in 4, 4, 2, and 2 comparisons, respectively. The group with incomplete evidence or no higher-certainty comparisons comprised local shockwave and spinal manual therapy, which lacked QoL data at all timepoints. Steroids and other injection therapies were also placed in this group because no comparison reached high or moderate of superiority with high certainty of evidence. The group with very limited evidence included ultrasound, laser, and taping, for which data were available only in the short-term. Needling was also classified in this group because mid- and long-term data were available only for function. Although evidence was available, education was rarely used as a sole treatment and sensitivity analyses showed reduced efficacy when restricted to single-focus education-only trials. Therefore, it could not be included into any of the other groups.

Sensitivity analyses restricted to interventions with a single treatment focus included fewer studies (short = 89, mid = 28, long = 11), resulting in fewer available comparisons with the minimal-intervention benchmark (Supplementary S18). Additional sensitivity analyses including only interventions with a single focus and no adjunct treatments included 36 studies in the short term and 31 in the mid-term, with insufficient data for long-term comparisons (Supplementary S19). Across both sensitivity analyses, results were largely consistent with the main analyses. Effect sizes and probabilities of superiority showed only minor variation, without a consistent directional shift. Notable exceptions were education-only and range-of-motion-only interventions, which demonstrated reduced efficacy when restricted to a single focus compared with analyses allowing multiple foci.

**Table 5.**
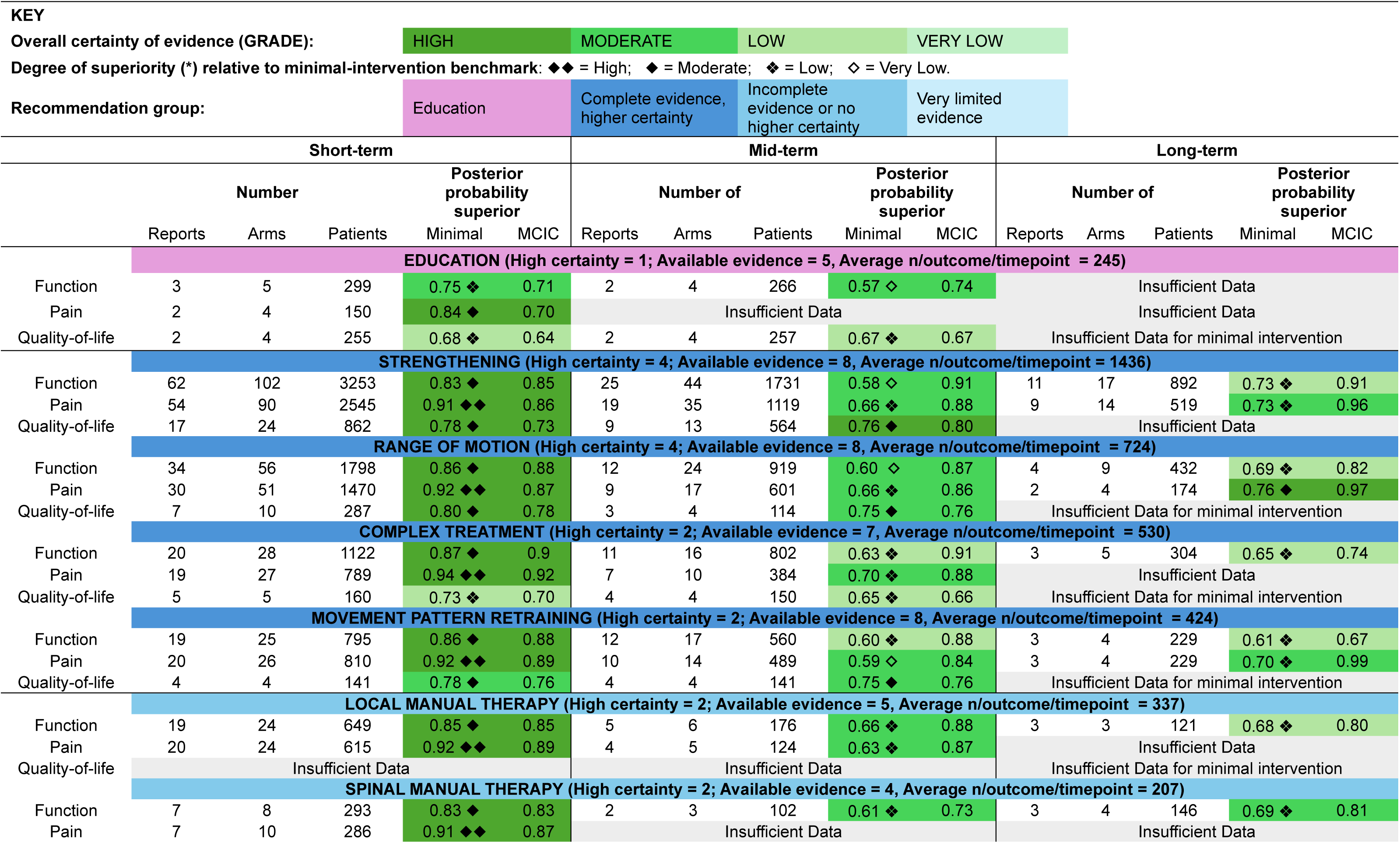
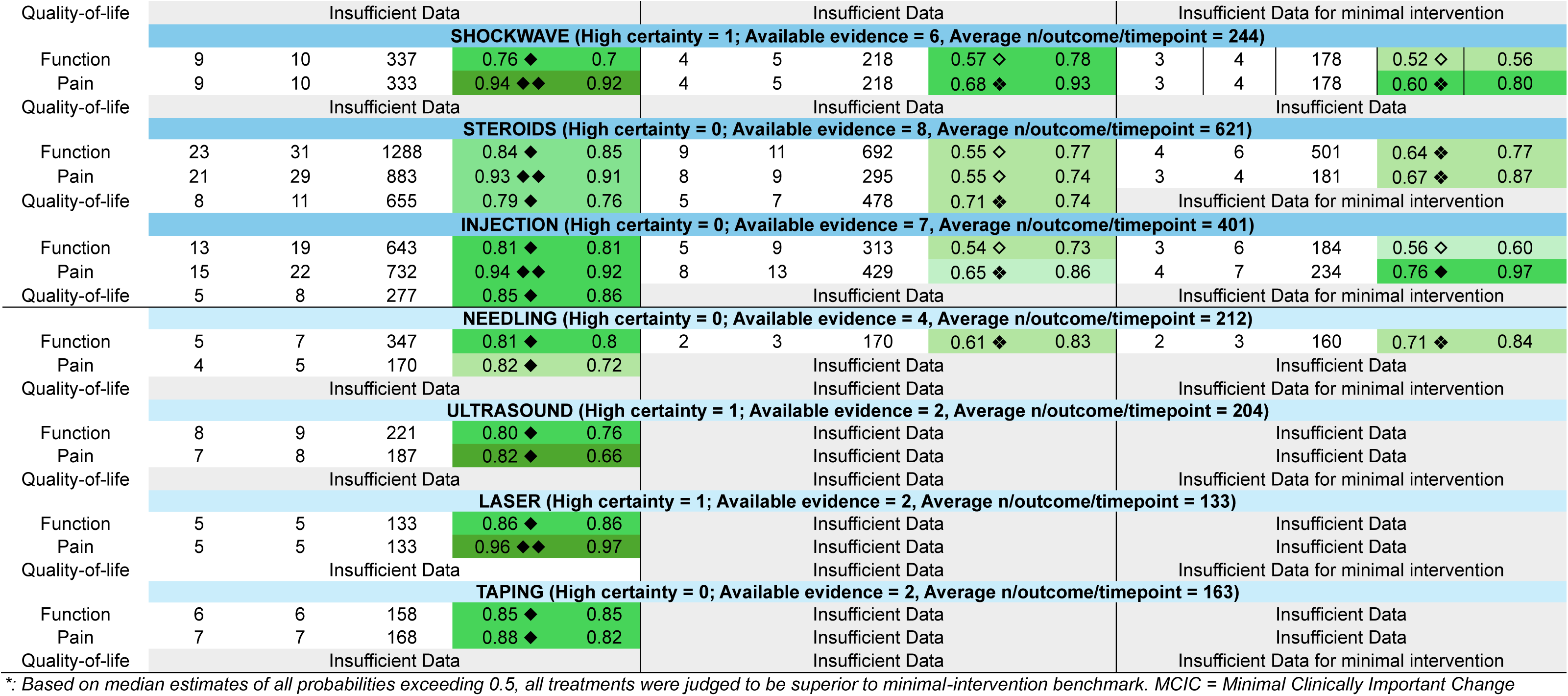
– Predictive meta-analysis results for pooled treatment categories, including probabilities of superiority relative to the minimal-intervention benchmark, probabilities of exceeding minimal clinically important change thresholds, and certainty of evidence.

### Network meta-analysis

Relative treatment effects informed jointly by direct and indirect evidence were synthesised using network meta-analyses restricted to single-focus interventions, minimal interventions (control), and interventions classified as complex (Figure 4 and Supplementary S20). Sufficient data and network connectivity were available to estimate relative effects for all outcomes in the short term, and for function in the mid and long term. Very serious imprecision was identified across all analyses, reflected by unstable ranking estimates in which many interventions spanned most possible rank positions based on 95% credible intervals (Figure 4). As a result, the overall certainty of evidence was judged as low for all assessments. Detailed results, including the number of contributing studies, study arms and participants, relative treatment effects compared with strengthening, SUCRA values, and full GRADE judgements, are provided in the Supplementary Materials.

**Figure 4.**
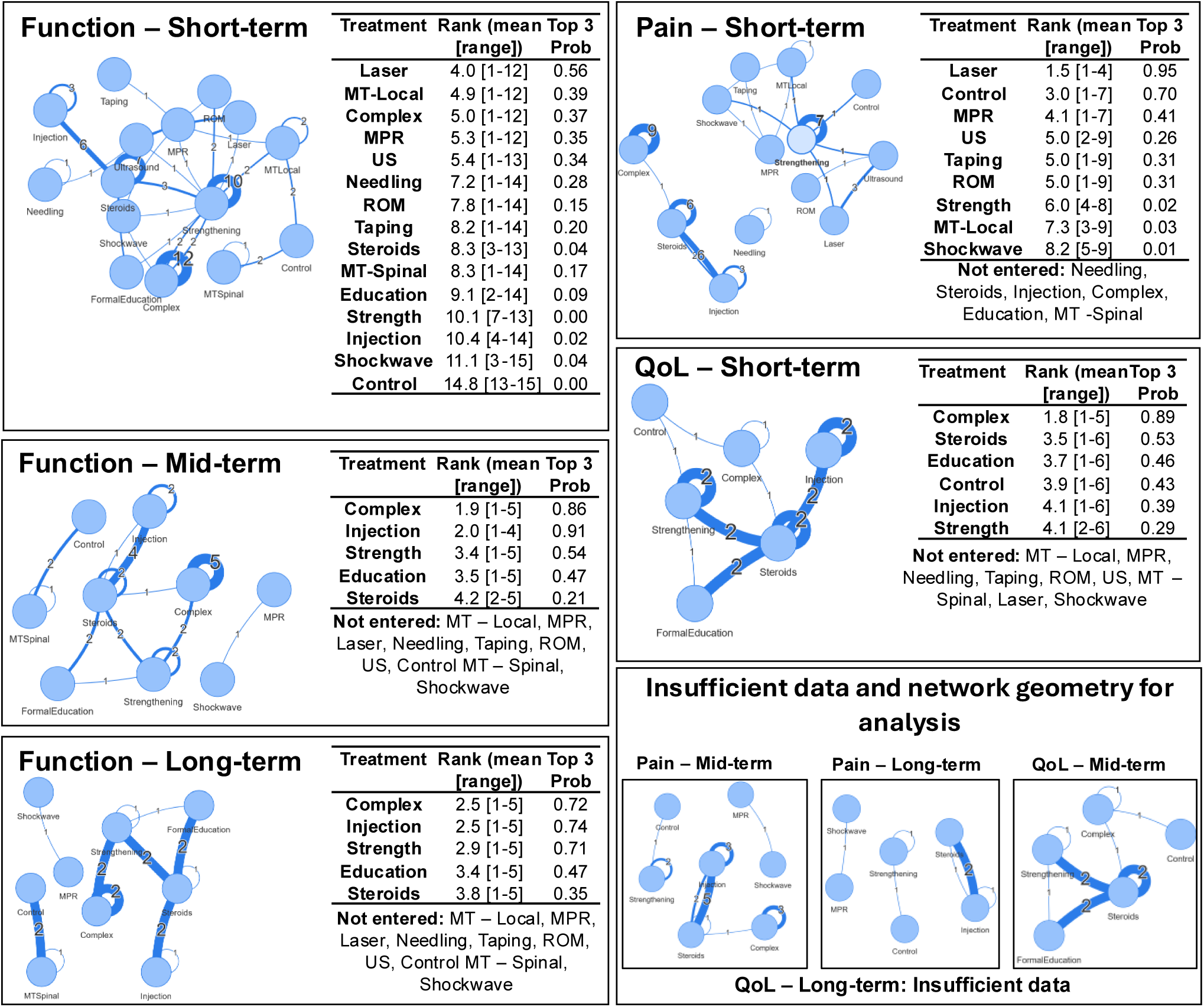
Network diagrams of RCT direct-comparison study arms restricted to minimal interventions, single-focus interventions, and interventions classified as complex, with corresponding network outputs including treatment rankings and probabilities of being among the top three treatments. Nodes represent treatments, and the number and thickness of edges indicate the number of direct head-to-head comparisons contributing to the network. For each treatment, the posterior mean rank and its range based on the 95% credible interval are shown, along with the posterior probability of being among the top three ranked treatments. Treatment ranks were derived from posterior samples of relative treatment effects, with higher ranks assigned to more favourable effects for the outcome of interest. QoL = Quality-of-life, MT = Manual Therapy, MPR = Movement Pattern Retraining, ROM = Range of Motion, US = Ultrasound

## DISCUSSION

We aimed to determine which treatments for RoCuTe are superior to minimal intervention and an identifiable hierarchy. Our findings demonstrate that all active treatment options included in the primary analyses, including complex interventions, were superior to minimal intervention for pain, function, and QoL across timepoints where there was sufficient data, but with substantial uncertainty for many comparisons, particularly beyond the short term. Recommendations were determined by integrating the GRADE-determined certainty of evidence with the consistency of superiority to minimal intervention across all timepoints and outcomes, showing that strengthening, range-of-motion exercises, complex interventions, and movement pattern retraining were supported by the most consistent and higher-certainty evidence across outcomes and timepoints. Clinicians should therefore implement these actives treatment at an early stage, with specifics indicated by clinical reasoning and patient preferences.

Despite the limited data available in some outcome-timepoint combinations, there was a clear hierarchy between the four levels of minimal intervention, with a wait-and-see approach demonstrating the poorest results. This indicates that even in the absence of active treatments, contact with the treatment provider leads to improvements due to contextual factors.^25,26^ These have been proposed to lead to placebo effects through the *classical conditioning* model (where patients have a conditioned response in the absence of an active principle because they have been previously associated with it) or the *expectation model* (where the expectancy of effects due to other factors triggers neurobiological changes that improve symptoms) ^26^. Finally, the interventions that contained sham plus an active secondary intervention showed consistently better improvements than the other minimal intervention levels. These secondary interventions were usually advice to exercise or general information provision, which strengthens the hypothesis that exercise and improved understanding of the condition are fundamental components of recovery from RoCuTe.^27,28^

Strengthening exercise was the most common treatment in this review, with 102 arms for function in the short-term and 17 in the long-term. As in clinical practice, studies in this review often combined strengthening with range-of-motion and movement pattern retraining exercises. This is likely due to exercise being recognised as the most common approach in clinical practice, as it can be readily prescribed with easy-to-obtain materials such as elastic bands and weights, be executed at home and improve patient self-efficacy.^29^ The consistent superiority of these treatments to minimal intervention across timepoints and outcomes may be primarily justified by the large number of studies. The causal mechanisms of exercise for RoCuTe can be classified into four domains: changes in tendon structure, neuromuscular performance, processing of pain and sensorimotor input and psychological factors.^30^ These lead to improvements in load capacity, flexibility, biomechanics and pain tolerance, ultimately decreasing pain and increasing function and quality of life.

RoCuTe interventions, both in clinical practice and in research, often comprise multiple treatments to address different components of the presentation such as load tolerance, knowledge of condition, self-efficacy, adherence, restricted range-of-motion, adverse movement patterns, pain beliefs and spinal elements; all while trying to also deal with the structural problem. Existing reviews have often classified interventions into a single treatment category, despite patients often receiving another treatment of a comparable dose.^12^ Our method of identifying multiple treatment foci and considering a group of “Complex Interventions” allowed us to assess efficacy of interventions that emulate clinical practice. Although these interventions could have targeted each treatment element at a different aspect of the presentation, the thought process behind these choices was rarely reported, making synthesis of the rationale difficult. Complex interventions which were more explicit were classified as well-developed yet did not show larger effects than less well-developed interventions, plausibly because better-developed interventions are typically evaluated in larger, more rigorously conducted studies where effect estimates tend to be smaller.^31^

Two trials included a carefully developed education-only group^32,33^ that showed limited efficacy, however in clinical practice education is typically delivered alongside active interventions as the main way of addressing a range of psychosocial factors and optimising adherence. Education-focused interventions had short- and mid-term data available but showed the smallest short-term improvements in pain and function, suggesting limited benefit up to 12 weeks, particularly when restricted to trials in which education was the sole treatment focus. In contrast, education seemed a more potent treatment for QoL in both the short- and mid-term and for function in the mid-term. This could be attributed to psychosocial issues being particularly correlated with QoL.^34,35^

Manual therapy, injection therapies and shockwave had lesser evidence of efficacy across multiple timepoints. Steroid injections are one of the most popular treatments in clinical practice and were investigated by many high-quality studies (n=23). Although commonly used, there is evidence that the benefits are mainly short-term and may be damaging in the long-term.^36^ Our findings do not support this claim for RoCuTe, as improvements from baseline were still demonstrated in the long-term, remaining superior to minimal intervention. The presence of publication bias means that the apparently positive results for steroid injection must be interpreted with particular caution.

The network meta-analysis, including interventions where there was only one focus, had a limited number of available arms for comparison, showed no treatment was consistently superior to others across outcomes and timepoints, presenting a wide range of possible ranks, indicating greater uncertainty. This finding, alongside the lack of a clear hierarchy in the primary analysis, further emphasises the need to deploy expert clinical reasoning and consider patient preferences when selecting treatments - choices being best guided by the primary analysis of efficacy.

### Limitations

To manage the volume and complexity of the available evidence, several methodological decisions were necessary and should be considered when interpreting our findings. Although many studies included mid- (>3 months) and long-term (>12 months) follow-up assessments, treatment duration itself was typically much shorter, averaging approximately six weeks. In some trials, only a single treatment session was delivered,^37–39^ while others did not clearly report duration or varied duration across treatment components.^40,41^ Treatment duration was not modelled as a moderator; its effects are therefore absorbed into residual error and between-study heterogeneity, and may contribute to uncertainty in mid- and long-term estimates.

Despite identifying 140 high-quality studies, each statistical comparison required alignment of outcome, timepoint, and treatment category, reducing the number of studies contributing to individual analyses. As a result, some comparisons were informed by relatively few studies and were therefore more sensitive to the influence of individual trials. This highlights the need for additional high-quality RCTs examining specific treatments (such as laser, ultrasound and taping), particularly with longer follow-up periods. A further limitation relates to the estimation of the minimal-intervention benchmark. For some outcome-timepoint combinations, there were insufficient wait-and-see data alone, requiring inclusion of higher-level minimal interventions (e.g., sham with or without secondary intervention). This likely inflated the estimated minimal-intervention change, thereby reducing the estimated probability of superiority for active treatments and making our conclusions conservative.

Importantly, although treatment categories were defined by a clear intervention focus, many included studies contained multiple treatment components. In the primary analysis, all studies containing a given focus were included in that category regardless of other foci and adjuncts, limiting causal attribution to any single component. However, sensitivity analyses restricted to single-focus interventions produced largely similar results, supporting the robustness of the main findings (Supplementary S18-S19). From a network perspective, analyses were restricted to control, single-focus and complex interventions to avoid structural overlap and cycles introduced by multi-component categorisation. Although this improved interpretability, it reduced the number of direct head-to-head comparisons, resulting in sparse network geometry, wide credible intervals, and unstable treatment rankings. This limits confidence in comparative hierarchies and underscores the need for more direct comparisons between clearly defined active interventions.

### Clinical application of findings

Our findings primarily suggest that doing something is far superior to doing nothing when managing a patient with RoCuTe. How superior depends on the outcome of interest and if the goals are for short- or long-term improvements. Overall, strengthening, range-of-motion and movement pattern retraining exercises as well as complex interventions showed the strongest profile (most data available with highest certainty of evidence) and are highly recommended. These findings are useful for clinicians, as not all treatments will be at their disposal, and some patients may not respond well to the initial intervention choice. Clinicians can therefore select and combine treatments based on their reasoning, availability, technique proficiency, and patient preferences.^42^ The findings of this meta-analysis can be combined with other research available describing experts’ clinical reasoning and patients’ experiences in order to find the optimal intervention approach^18,43^

## CONCLUSIONS

This review of high-quality studies on RoCuTe management provides strong and consistent evidence that active treatments are superior to minimal interventions. Clinicians should prioritise strengthening, range-of-motion exercises, movement pattern retraining, and complex treatment combinations when selecting active management, as they have the most consistently supported evidence across outcomes and timepoints where comparative data are available. The findings of this review should be combined with expert clinical reasoning alongside patient preferences in selecting intervention content and timing.

## STATEMENTS

## Data Availability

All data produced in the present study are available upon reasonable request to the authors

## Acknowledgements

We would like to particularly acknowledge the patients, public, professionals and other stakeholders who contributed to the PPIEP activities and participated in this study. We would also like to acknowledge the funders for their unwavering support, QMUL colleagues in the joint research management office, and Gill Morrey for project management.

## Role of the funding source

This work was supported by the Private Physiotherapy Educational Foundation Silver Jubilee Award (QMUL reference 13569244).

This work acknowledges the support of the National Institute for Health Research Barts Biomedical Research Centre (NIHR203330) for researchers employed at QMUL and Barts Health NHS trust.

This work acknowledges the support of Podium Analytics and the Podium Institute, a charity with the mission of More Sport Less Injury. https://podiumanalytics.org

The funders had no role in considering the study design, in the collection, analysis, or interpretation of data or in the writing of the report.

## Ethical approval

Not required

## Data sharing

Data can be made available upon request to the PI, Professor Morrissey.

## Patient and public involvement

Patients and/or the public were involved in the design, conduct, and reporting of this research. Refer to the methods section for further details.

## Dissemination to participants and related patient and public communities

We plan to disseminate the findings of this study to lay audiences through mainstream and social media. Additionally, this will inform a best practice guide (in combination with clinician reasoning and patient perspectives qualitative studies) which will form the basis of an education and international implementation strategy.

## Transparency statement

The study guarantor (DM) affirms that the manuscript is an honest, accurate, and transparent account of the study being reported; that no important aspects of the study have been omitted; and that any discrepancies from the study as planned (and, if relevant, registered) have been explained.

