## Supplementary material for "Active treatments outperform minimal intervention for adults with rotator cuff tendinopathy: a systematic review with predictive and network meta-analyses of complex interventions": SOut.html

SOut.knit


### Title: Systematic review with Bayesian predictive meta-analysis and network meta-analysis

##### Supplementary file contents

- S1. Prisma checklist - Search (open
  PDF)
- S2. Complex interventions and NMA Prisma
  checklist (open
  PDF)
- S3. Operational definitions of the terms
  used (open
  PDF)
- S4. Search terms (open
  PDF)
- S5. Search results (open
  PDF)
- S6. Extracted items (open
  PDF)
- S7. Strengthening
  classification (open
  PDF)
- S8. General classification
  (open
  PDF)
- S9. Complex classification
  (open
  PDF)
- S10. Category coding (open
  PDF)
- S11. Minimal-intervention benchmarking (model
  results)
- S12. Publication bias (open
  PDF)
- S13. List of excluded studies (open
  PDF)
- S14. List of low-quality studies
  (open
  PDF)
- S15. Mimial-intervention studies
  (open
  PDF)
- S16. Active analysis (model results)
- S17. Active-intervention studies
  (open
  PDF)
- S18. Active analysis sensitivity 1 (model
  results)
- S19. Active analysis sensitivity 2 (model
  results)

#### S1. Prisma checklist - Search

Embedded PDF not supported.
Download
S1 PDF

#### S2. Complex Intervention and NMA Prisma checklist

Embedded PDF not supported.
Download
S2 PDF

#### S3. Operational definitions of the terms used

Embedded PDF not supported.
Download
S3 PDF

#### S4. Search terms

Embedded PDF not supported.
Download
S4 PDF

#### S5. Search results

Embedded PDF not supported.
Download
S5 PDF

#### S6. Extracted items

Embedded PDF not supported.
Download
S6 PDF

#### S7. Strengthening classification

Embedded PDF not supported.
Download
S7 PDF

#### S8. General classification

Embedded PDF not supported.
Download
S8 PDF

#### S9. Complex classification

Embedded PDF not supported.
Download
S9 PDF

#### S10. Category coding

Embedded PDF not supported.
Download
S10 PDF

#### S11.Bayesian predictive meta-analyses of minimal intervention categories

##### Read me

**What this file contains.** Summary tables and figures for
Minimal Intervention analyses, organised by Outcome (Pain / Function /
QoL) and Time (Short / Mid / Long).

- **Index (below):** Use the global search (top-right of the
  table) or the per-column filters to find an item (e.g., type an author
  surname in *Studies*). Click *View* to jump to the
  section.
- **Results tabs:** The *Results* area is grouped into
  tabs: Tables – Comparison, Tables – Benchmark, Figures – Comparison, and
  Figures – Benchmark.
- **Anchors:** Each section header has an anchor (e.g.,
  `Pain — Short`). The Index’s *View* links scroll
  directly to these.
- **Missing files:** If a saved file isn’t present alongside
  this HTML, you’ll see a “Missing file” message in its place.
- **Print / PDF:** Use your browser’s `Ctrl/Cmd +
  P`. For widest tables, prefer landscape orientation.

#### Index (Global search + column filters)

### Results

#### Tables Minimal Intervention Comparison

###### ***Notes for table: Meta-analyses were performed across minimal intervention classes (wait-and-see, inert sham, active sham, sham plus plausible secondary intervention) when ≥2 studies were available. n\_Total is the total number of participants across included studies. Effect is the posterior median pooled effect size with its 75% credible interval (CrI). SUCRA (Surface Under the Cumulative Ranking Curve) summarises, across the posterior predictive distributions the probability that a given minimal intervention class yields the largest effect (range 0–1; higher = more likely best).***

##### Pain — Short

| Studies | Intervention | Outcome | Duration | n\_Studies | n\_Effects | n\_Total | Effect | SUCRA |
| --- | --- | --- | --- | --- | --- | --- | --- | --- |
| Lombardi et al. 2008, Eliason et al. 2021, Letafatkar et al. 2021 | Wait-and-See | Pain | Short | 3 | 3 | 109 | -2.57 (-8.73 to 3.57) | 0.16 |
| Hunter et al. 2022, Taik et al. 2022, Bennell et al. 2010, Li et al. 2017, Kolk et al. 2013, Atya 2012, Munday et al. 2007, Shakeri et al. 2013, Speed et al. 2002, Vecchio et al. 1993 | Inert Sham | Pain | Short | 10 | 10 | 298 | -13.51 (-17.10 to -9.92) | 0.46 |
| Delgado-Gil et al. 2015, Chou et al. 2010, Barış Bayram et al. 2014, Itzkowitch et al. 1996, Moghtaderi et al. 2013, Rueda Garrido et al. 2016 | Active Sham | Pain | Short | 6 | 6 | 184 | -17.53 (-20.35 to -14.53) | 0.62 |
| Nazligul et al. 2018, Sandford et al. 2018, Kelle et al. 2023, Paoloni et al. 2005, Szczurko et al. 2009, Paavola et al. 2018, Kocyigit et al. 2016, Akbaş et al. 2025, Baeske et al. 2024, Karasu et al. 2024, Kim et al. 2025 | Sham + Secondary | Pain | Short | 11 | 12 | 356 | -23.17 (-27.86 to -18.41) | 0.76 |

##### Pain — Mid

| Studies | Intervention | Outcome | Duration | n\_Studies | n\_Effects | n\_Total | Effect | SUCRA |
| --- | --- | --- | --- | --- | --- | --- | --- | --- |
| Eliason et al. 2021 | Wait-and-See | Pain | Mid | 1 | 1 | 39 | — | — |
| Hunter et al. 2022, Bennell et al. 2010, Kolk et al. 2013, Brox et al. 1999, Speed et al. 2002 | Inert Sham | Pain | Mid | 5 | 5 | 177 | -20.17 (-28.45 to -12.27) | 0.32 |
| — | Active Sham | Pain | Mid | — | — | — | — | — |
| Sandford et al. 2018, Kelle et al. 2023, Paoloni et al. 2005, Paavola et al. 2018 | Sham + Secondary | Pain | Mid | 4 | 5 | 147 | -27.71 (-30.24 to -25.11) | 0.68 |

##### Pain — Long

| Studies | Intervention | Outcome | Duration | n\_Studies | n\_Effects | n\_Total | Effect | SUCRA |
| --- | --- | --- | --- | --- | --- | --- | --- | --- |
| — | Wait-and-See | Pain | Long | — | — | — | — | — |
| Hunter et al. 2022, Brox et al. 1999 | Inert Sham | Pain | Long | 2 | 2 | 38 | -21.13 (-37.68 to -5.56) | 0.26 |
| — | Active Sham | Pain | Long | — | — | — | — | — |
| Sandford et al. 2018, Paavola et al. 2018 | Sham + Secondary | Pain | Long | 2 | 3 | 98 | -40.64 (-44.39 to -36.51) | 0.74 |

##### Function — Short

| Studies | Intervention | Outcome | Duration | n\_Studies | n\_Effects | n\_Total | Effect | SUCRA |
| --- | --- | --- | --- | --- | --- | --- | --- | --- |
| Lombardi et al. 2008, Eliason et al. 2021, Letafatkar et al. 2021, Ludewig & Borstad 2003, Khandaloo et al. 2025 | Wait-and-See | Function | Short | 5 | 5 | 159 | 0.13 (-0.30 to 0.57) | 0.24 |
| Hunter et al. 2022, Taik et al. 2022, Bennell et al. 2010, Li et al. 2017, Kolk et al. 2013, Atya 2012, Kleinhenz et al. 1999, Speed et al. 2002 | Inert Sham | Function | Short | 8 | 9 | 279 | 0.73 (0.55 to 0.90) | 0.5 |
| Chou et al. 2010, Moghtaderi et al. 2013, Rueda Garrido et al. 2016 | Active Sham | Function | Short | 3 | 3 | 79 | 0.77 (-0.41 to 1.54) | 0.49 |
| Nazligul et al. 2018, Belley et al. 2018, Sandford et al. 2018, Kelle et al. 2023, Szczurko et al. 2009, Kocyigit et al. 2016, Akbaş et al. 2025, Baeske et al. 2024, Karasu et al. 2024, Kim et al. 2025 | Sham + Secondary | Function | Short | 10 | 12 | 286 | 1.35 (1.02 to 1.71) | 0.77 |

##### Function — Mid

| Studies | Intervention | Outcome | Duration | n\_Studies | n\_Effects | n\_Total | Effect | SUCRA |
| --- | --- | --- | --- | --- | --- | --- | --- | --- |
| Dickens et al. 2005, Eliason et al. 2021 | Wait-and-See | Function | Mid | 2 | 2 | 79 | 0.87 (-0.65 to 2.36) | 0.42 |
| Hunter et al. 2022, Bennell et al. 2010, Kolk et al. 2013, Brox et al. 1999, Speed et al. 2002 | Inert Sham | Function | Mid | 5 | 6 | 177 | 1.04 (0.81 to 1.27) | 0.43 |
| — | Active Sham | Function | Mid | — | — | — | — | — |
| Sandford et al. 2018, Kelle et al. 2023, Paavola et al. 2018 | Sham + Secondary | Function | Mid | 3 | 4 | 120 | 1.79 (0.87 to 2.73) | 0.65 |

##### Function — Long

| Studies | Intervention | Outcome | Duration | n\_Studies | n\_Effects | n\_Total | Effect | SUCRA |
| --- | --- | --- | --- | --- | --- | --- | --- | --- |
| — | Wait-and-See | Function | Long | — | — | — | — | — |
| Hunter et al. 2022, Brox et al. 1999 | Inert Sham | Function | Long | 2 | 3 | 38 | 0.76 (-0.05 to 1.62) | 0.33 |
| — | Active Sham | Function | Long | — | — | — | — | — |
| Sandford et al. 2018, Paavola et al. 2018 | Sham + Secondary | Function | Long | 2 | 3 | 98 | 1.92 (0.66 to 3.08) | 0.67 |

##### QoL — Short

| Studies | Intervention | Outcome | Duration | n\_Studies | n\_Effects | n\_Total | Effect | SUCRA |
| --- | --- | --- | --- | --- | --- | --- | --- | --- |
| Lombardi et al. 2008 | Wait-and-See | QoL | Short | 1 | 8 | 30 | — | — |
| Bennell et al. 2010 | Inert Sham | QoL | Short | 1 | 2 | 61 | — | — |
| — | Active Sham | QoL | Short | — | — | — | — | — |
| Belley et al. 2018, Sandford et al. 2018, Kelle et al. 2023, Szczurko et al. 2009, Kocyigit et al. 2016, Kim et al. 2025 | Sham + Secondary | QoL | Short | 6 | 17 | 165 | 0.23 (0.02 to 0.49) | — |

##### QoL — Mid

| Studies | Intervention | Outcome | Duration | n\_Studies | n\_Effects | n\_Total | Effect | SUCRA |
| --- | --- | --- | --- | --- | --- | --- | --- | --- |
| — | Wait-and-See | QoL | Mid | — | — | — | — | — |
| Bennell et al. 2010 | Inert Sham | QoL | Mid | 1 | 2 | 61 | — | — |
| — | Active Sham | QoL | Mid | — | — | — | — | — |
| Sandford et al. 2018, Kelle et al. 2023 | Sham + Secondary | QoL | Mid | 2 | 7 | 57 | 0.43 (-0.24 to 1.03) | — |

##### QoL — Long

| Studies | Intervention | Outcome | Duration | n\_Studies | n\_Effects | n\_Total | Effect | SUCRA |
| --- | --- | --- | --- | --- | --- | --- | --- | --- |
| — | Wait-and-See | QoL | Long | — | — | — | — | — |
| — | Inert Sham | QoL | Long | — | — | — | — | — |
| — | Active Sham | QoL | Long | — | — | — | — | — |
| Sandford et al. 2018 | Sham + Secondary | QoL | Long | 1 | 1 | 35 | — | — |

#### Tables Minimal Intervention Benchmark

###### ***Notes for table: Benchmark meta-analyses were performed for each time period (short, mid, long) when ≥2 studies were available. Evidence was added hierarchically to meet this criteria. First with wait-and-see studies, then combining or using inert sham, and up to sham plus plausible secondary intervention if required. n\_Total is the total number of participants across included studies. Effect is the posterior median pooled effect size with its 50% credible interval (CrI). SUCRA (Surface Under the Cumulative Ranking Curve) summarises, across the posterior predictive distributions for the three time periods, the probability that a given period yields the largest effect (range 0–1; higher = more likely best).***

##### Pain — Short/Mid/Long

| Studies | Intervention | Outcome | Duration | n\_Studies | n\_Effects | n\_Total | Effect | SUCRA |
| --- | --- | --- | --- | --- | --- | --- | --- | --- |
| Lombardi et al. 2008, Eliason et al. 2021, Letafatkar et al. 2021 | Wait-and-See:3 | Pain | Short | 3 | 3 | 109 | -2.57 (-8.73 to 3.57) | 0.19 |
| Eliason et al. 2021, Hunter et al. 2022, Bennell et al. 2010, Kolk et al. 2013, Brox et al. 1999, Speed et al. 2002 | Wait-and-See:1, Inert Sham:5 | Pain | Mid | 6 | 6 | 216 | -23.34 (-29.25 to -17.09) | 0.71 |
| Hunter et al. 2022, Brox et al. 1999 | Inert Sham:2 | Pain | Long | 2 | 2 | 38 | -21.13 (-37.68 to -5.56) | 0.61 |

##### Function — Short/Mid/Long

| Studies | Intervention | Outcome | Duration | n\_Studies | n\_Effects | n\_Total | Effect | SUCRA |
| --- | --- | --- | --- | --- | --- | --- | --- | --- |
| Lombardi et al. 2008, Eliason et al. 2021, Letafatkar et al. 2021, Ludewig & Borstad 2003, Khandaloo et al. 2025 | Wait-and-See:5 | Function | Short | 5 | 5 | 159 | 0.13 (-0.30 to 0.57) | 0.37 |
| Dickens et al. 2005, Eliason et al. 2021 | Wait-and-See:2 | Function | Mid | 2 | 2 | 79 | 0.87 (-0.65 to 2.36) | 0.56 |
| Hunter et al. 2022, Brox et al. 1999 | Inert Sham:2 | Function | Long | 2 | 3 | 38 | 0.76 (-0.05 to 1.62) | 0.57 |

##### QoL — Short/Mid/Long

| Studies | Intervention | Outcome | Duration | n\_Studies | n\_Effects | n\_Total | Effect | SUCRA |
| --- | --- | --- | --- | --- | --- | --- | --- | --- |
| Lombardi et al. 2008, Bennell et al. 2010 | Wait-and-See:1, Inert Sham:1 | QoL | Short | 2 | 10 | 91 | 0.05 (-0.37 to 0.47) | 0.33 |
| Sandford et al. 2018, Kelle et al. 2023, Bennell et al. 2010 | Inert Sham:1, Sham + Secondary:2 | Q0L | Mid | 3 | 9 | 118 | 0.33 (0.02 to 0.63) | 0.67 |
| Sandford et al. 2018 | Sham + Secondary:1 | QoL | Long | 1 | 1 | 35 | — | — |

#### Figures Minimal Intervention Comparison

###### ***Notes for figures: Plots are based on meta-analyses across minimal-intervention classes (wait-and-see, inert sham, active sham, sham plus plausible secondary intervention) and are shown only when ≥2 studies were available. Each plot displays the posterior predictive distribution of effects for future studies; the blue band marks the middle 50% (interquartile) of that distribution.***

##### Pain — Short

##### Pain — Mid

##### Pain — Long

##### Function — Short

##### Function — Mid

##### Function — Long

##### QoL — Short

##### QoL — Mid

##### QoL — Long

###### ***Insufficient Data***

#### Figures Minimal Intervention Benchmark

###### ***Notes for figures: Plots are based on benchmark meta-analyses that were performed for each time period (short, mid, long) when ≥2 studies were available. Evidence was added hierarchically to meet this criteria. First with wait-and-see studies, then combining or using inert sham, and up to sham plus plausible secondary intervention if required. Each plot displays the posterior predictive distribution of effects for future studies; the blue band marks the middle 50% (interquartile) of that distribution.***

##### Pain — Short/Mid/Long

##### Function — Short/Mid/Long

##### QoL — Short/Mid/Long

#### S12. Publication bias

Embedded PDF not supported.
Download
S12 PDF

#### S13. List of excluded studies

Embedded PDF not supported.
Download
S13 PDF

#### S14. List of low-quality studies

Embedded PDF not supported.
Download
S14 PDF

#### S15. Minimal-intervention study characteristics

Embedded PDF not supported.
Download
S15 PDF

#### S16. Bayesian predictive meta-analyses comparing active interventions with minimal-intervention benchmarking, and network meta-analyses comparing active interventions

### Outcomes

#### Pain

##### Short

###### Mapping

**Figure 1. Focus map without adjunct interventions**  
This figure shows the distribution of the intervention modalities
identified as a focus and their
overlaps.  
  
  
**Figure
2. Focus map including adjunct interventions**  
 This figure
shows the distribution of the intervention modalities identified as a
focus, the adjuncts, and their
overlaps.  
  
  
**Table
1. Summary of interventions**  
 This table summarises the
frequency of intervention modalities identified as a focus across
studies, study arms, and the number of participants included.

```
##         Intervention Studies Groups TotalN
## 1      Strengthening      54     90   2545
## 2                ROM      30     51   1470
## 3           Steroids      21     29    883
## 4                MPR      20     26    810
## 5            MTLocal      20     24    615
## 6   ComplexTreatment      19     27    789
## 7          Injection      15     22    732
## 8          Shockwave       9     10    333
## 9           MTSpinal       7     10    286
## 10        Ultrasound       7      8    187
## 11            Taping       7      7    168
## 12             Laser       5      5    133
## 13          Needling       4      5    170
## 14      Electrolysis       4      4     85
## 15   FormalEducation       2      4    150
## 16              TENS       2      4     95
## 17    Radiofrequency       2      3     81
## 18 Ultrasonophoresis       2      3     57
## 19         Diathermy       2      2     48
## 20               IFC       2      2     45
## 21           Thermal       2      2     37
## 22   Electromagnetic       1      1     40
## 23              NMES       1      1     20
```

###### Summary Table

###### GRADE Table

GRADE Summary (Pain Short-Term)

| Intervention | total\_N | Risk of bias | Imprecision | Inconsistency | Indirectness | Publication bias |
| --- | --- | --- | --- | --- | --- | --- |
| Laser | 133 | No serious risk of bias | No serious imprecision | No serious inconsistency | No serious indirectness | Undetected |
| Injection | 732 | No serious risk of bias | No serious imprecision | No serious inconsistency | No serious indirectness | Strongly suspected |
| Steroids | 883 | No serious risk of bias | No serious imprecision | No serious inconsistency | No serious indirectness | Strongly suspected |
| Complex | 789 | No serious risk of bias | No serious imprecision | No serious inconsistency | No serious indirectness | Undetected |
| Shockwave | 333 | No serious risk of bias | No serious imprecision | No serious inconsistency | No serious indirectness | Undetected |
| MT-Local | 615 | No serious risk of bias | No serious imprecision | No serious inconsistency | No serious indirectness | Undetected |
| MT-Spinal | 286 | No serious risk of bias | No serious imprecision | No serious inconsistency | No serious indirectness | Undetected |
| ROM | 1470 | No serious risk of bias | No serious imprecision | No serious inconsistency | No serious indirectness | Undetected |
| Strengthening | 2545 | No serious risk of bias | No serious imprecision | No serious inconsistency | No serious indirectness | Undetected |
| MPR | 810 | No serious risk of bias | No serious imprecision | No serious inconsistency | No serious indirectness | Undetected |
| Taping | 168 | No serious risk of bias | No serious imprecision | Serious inconsistency | No serious indirectness | Undetected |
| Needling | 170 | No serious risk of bias | No serious imprecision | Serious inconsistency | No serious indirectness | Strongly suspected |
| Ultrasound | 187 | No serious risk of bias | No serious imprecision | No serious inconsistency | No serious indirectness | Undetected |
| Education | 150 | No serious risk of bias | No serious imprecision | No serious inconsistency | No serious indirectness | Undetected |

###### Summary Figure

**Figure 1. Effect size plots** Shows the
minimal‐intervention benchmark alongside interventions that included
specific modalities as a primary focus (either alone or as one of
several components). The blue band represents the median and 75%
credible interval for the pooled meta-analytic mean. The shaded
distribution illustrates the predicted effect sizes expected from a
future study using a focused intervention. Black lines indicate the 66%
and 95% middle credible intervals of this predictive
distribution.

###### Risk of bias

Risk of Bias (Pain Short-Term)

| Study | RoB |
| --- | --- |
| Arias-Buría et al. 2017 | LOW |
| Østerås et al. 2010 | LOW |
| Tauqeer et al. 2024 | LOW |
| Santamato et al. 2009a | LOW |
| Marzetti et al. 2014 | LOW |
| Camargo et al. 2015 | LOW |
| Struyf et al. 2013 | LOW |
| Ehsani et al. 2024 | LOW |
| Eliason et al. 2021 | LOW |
| Kara et al. 2024 | LOW |
| Letafatkar et al. 2021 | LOW |
| Gomes et al. 2018 | LOW |
| Avendaño-Coy et al. 2022 | HIGH |
| de Oliveira et al. 2022 | LOW |
| Abu EL Kasem et al. 2024 | LOW |
| Pekgöz et al. 2020 | LOW |
| Schydlowsky et al. 2022 | LOW |
| Dupuis et al. 2018 | LOW |
| Eliason et al. 2022 | LOW |
| Raeesi et al. 2023 | LOW |
| Dubé et al. 2023 | LOW |
| Dejaco et al. 2017 | SOME |
| Rodríguez-Huguet et al. 2020 | HIGH |
| Kandemir et al. 2024 | SOME |
| Gutiérrez-Espinoza et al. 2023a | LOW |
| Liu et al. 2024 | LOW |
| Beltrán et al. 2024 | LOW |
| Hotta et al. 2020 | LOW |
| Ketola et al. 2009 | HIGH |
| Dilek et al. 2016 | LOW |
| Li et al. 2024 | LOW |
| Sen et al. 2023 | LOW |
| Türksan et al. 2024 | LOW |
| San Segundo et al. 2008 | LOW |
| Kesikburun et al. 2013 | LOW |
| Juul-Kristensen et al. 2019 | LOW |
| Lombardi et al. 2008 | LOW |
| Kulakli et al. 2020 | LOW |
| Kamonseki et al. 2023 | LOW |
| Nguyen et al. 2024 | LOW |
| Gutiérrez Espinoza et al. 2023b | LOW |
| Góngora-Rodríguez et al. 2024 | LOW |
| Paavola et al. 2018 | LOW |
| Mulligan et al. 2016 | LOW |
| Ingwersen et al. 2017 | HIGH |
| de Miguel Valtierra et al. 2018 | LOW |
| Arias-Buriá et al. 2015 | LOW |
| Bal et al. 2009 | LOW |
| Kromer et al. 2014 | LOW |
| Delgado-Gil et al. 2025 | LOW |
| Holmgren et al. 2012 | SOME |
| Buccioli et al. 2025 | LOW |
| Qureshi et al. 2024 | HIGH |
| Cavaggion et al. 2024 | LOW |
| Tahran & Yeşilyaprak 2020 | HIGH |
| Granviken & Vasseljen 2015 | HIGH |
| Szczurko et al. 2009 | LOW |
| Giombini et al. 2006 | HIGH |
| Babaei-Ghazani et al. 2019 | LOW |
| Ogbeivor et al. 2019 | LOW |
| Jo et al. 2020 | LOW |
| Dogu et al. 2012 | LOW |
| Turgut et al. 2025 | LOW |
| Lee et al. 2011 | LOW |
| Akbari et al. 2020 | LOW |
| Apivatgaroon et al. 2023 | LOW |
| Ebadi et al. 2023 | LOW |
| Boonard et al. 2018 | LOW |
| Penning et al. 2012 | LOW |
| Ekeberg et al. 2009 | LOW |
| Daghiani et al. 2023 | LOW |
| Wang et al. 2019 | LOW |
| Cole et al. 2016 | LOW |
| Cole et al. 2018 | LOW |
| Rossi et al. 2024 | SOME |
| Nazary-Moghadam et al. 2025 | SOME |
| de Oliveira et al. 2021 | SOME |
| Kim et al. 2020 | SOME |
| Doweir et al. 2025 | LOW |
| Engebretsen et al. 2009 | LOW |
| Dunning et al. 2021 | LOW |
| Delgado-Gil et al. 2015 | LOW |
| Ishaq et al. 2024 | LOW |
| Baeske et al. 2024 | LOW |
| Hunter et al. 2022 | LOW |
| Karimiahmadabadi et al. 2025 | LOW |
| Bennell et al. 2010 | LOW |
| Sırlan et al. 2025 | HIGH |
| Chou et al. 2010 | HIGH |
| Esmaily et al. 2022 | LOW |
| Ottani et al. 2024 | LOW |
| Ko et al. 2024 | LOW |
| Kim et al. 2025 | LOW |
| Li et al. 2017 | LOW |
| Kolk et al. 2013 | LOW |
| Kvalvaag et al. 2017 | LOW |
| Vinuesa-Montoya et al. 2017 | LOW |
| Yavuz et al. 2014 | SOME |
| Santamato et al. 2009b | LOW |
| Taik et al. 2022 | LOW |
| Kocyigit et al. 2016 | LOW |
| Taheri et al. 2021 | LOW |
| Kibar et al. 2017 | LOW |
| Akbaş et al. 2025 | LOW |
| Shin et al. 2021 | SOME |

###### Network Analysis

**Figure 1. Interactive network plot**  
 This figure
shows the pairwise comparisons between single-focus interventions that
were included in the corresponding benchmark analysis. Edge thickness
represents the number of pairwise comparisons contributing to each
connection. The nodes can be manipulated, with zooming and translation
using the mouse.

**Table 1. Network analysis results**  
 This table
summarises the mean pairwise difference in pain reduction (absolute
units: 0–100) between each single-focus intervention and the reference
treatment, strengthening. Positive values indicate that strengthening is
superior; negative values indicate that strengthening is
inferior.

**Table 2. Network ranking results**  
 This table
summarises the ranking of interventions from the network analysis,
including the mean rank and its uncertainty, the probability of ranking
1st and in the top 3, and the SUCRA (Surface Under the Cumulative
Ranking Curve) value with its uncertainty.  
  

| treatment | mean\_rank | rank\_lower | rank\_upperr | mean\_sucra | sucra\_lower | sucra\_upper | prob\_rank\_1 | prob\_rank\_1\_to\_3 |
| --- | --- | --- | --- | --- | --- | --- | --- | --- |
| Laser | 1.52 | 1 | 4 | 0.93 | 0.62 | 1.00 | 0.68 | 0.95 |
| Control | 2.97 | 1 | 7 | 0.75 | 0.25 | 1.00 | 0.17 | 0.70 |
| MPR | 4.09 | 1 | 7 | 0.61 | 0.25 | 1.00 | 0.06 | 0.41 |
| Ultrasound | 4.95 | 2 | 9 | 0.51 | 0.00 | 0.88 | 0.00 | 0.26 |
| Taping | 4.97 | 1 | 9 | 0.50 | 0.00 | 1.00 | 0.05 | 0.31 |
| ROM | 4.99 | 1 | 9 | 0.50 | 0.00 | 1.00 | 0.04 | 0.31 |
| Strengthening | 5.96 | 4 | 8 | 0.38 | 0.12 | 0.62 | 0.00 | 0.02 |
| MTLocal | 7.33 | 3 | 9 | 0.21 | 0.00 | 0.75 | 0.00 | 0.03 |
| Shockwave | 8.22 | 5 | 9 | 0.10 | 0.00 | 0.50 | 0.00 | 0.01 |

**Table 3. Network GRADE analysis**  
 This table
summarises the GRADE assessment of the network analysis.   
  

GRADE Summary of Findings (Pain, Short-Term)

| Analysis | total\_N | Risk of bias | Imprecision | Inconsistency | Indirectness | Publication bias |
| --- | --- | --- | --- | --- | --- | --- |
| Pain (short-term) | 585 | No serious risk of bias | Very serious imprecision | No serious inconsistency | No serious indirectness | Unclear |
|  |  | Most contributing studies were randomised trials with acceptable methods; risk of bias was unlikely to materially affect relative treatment comparisons. | Relative treatment effects were small compared with typical within-arm pain reductions, with wide credible intervals crossing no difference. Ranking estimates were unstable, with several interventions spanning most possible rank positions and wide SUCRA intervals. | Between-study heterogeneity was moderate-to-substantial (τ = 5.8; 95% CrI 5.0 to 8.3 on the pain-change scale). Global model fit was adequate (posterior mean residual deviance = 26.4 vs 26 arm-level observations). Allowing for inconsistency did not improve model fit (Base model mean Dres = 26.4, Inconsistency model mean Dres = 26.2, ΔDres = 0.2), indicating agreement between direct and indirect evidence. | Included studies directly addressed the population, interventions, and pain outcomes of interest; indirect comparisons were clinically plausible. | Formal assessment of small-study effects was limited by the number of studies contributing to each comparison. |

###### Mapping: Sensitivity Outcome

**Figure 1. Focus map without adjunct interventions**  
This figure shows the distribution of the intervention modalities
identified as a focus and their
overlaps.  
  
  
**Figure
2. Focus map including adjunct interventions**  
 This figure
shows the distribution of the intervention modalities identified as a
focus, the adjuncts, and their
overlaps.  
  
  
**Table
1. Summary of interventions**  
 This table summarises the
frequency of intervention modalities identified as a focus across
studies, study arms, and the number of participants included.

```
##            Intervention Studies Groups TotalN
## 1                   ROM      30     49   1440
## 2         Strengthening      30     46   1291
## 3              Steroids      21     29    883
## 4               MTLocal      20     24    615
## 5                   MPR      20     22    664
## 6         HighIntensity      15     24    756
## 7          LowIntensity      11     19    482
## 8             Shockwave       9     10    333
## 9              MTSpinal       7     10    286
## 10           Ultrasound       7      8    187
## 11               Taping       7      7    168
## 12                Laser       5      5    133
## 13          Hyaluroinic       4      7    223
## 14          Anaesthetic       4      6    255
## 15             Needling       4      5    170
## 16         Electrolysis       4      4     85
## 17                  PRP       3      3     95
## 18      FormalEducation       2      4    150
## 19                 TENS       2      4     95
## 20                Ozone       2      3     87
## 21       Radiofrequency       2      3     81
## 22    Ultrasonophoresis       2      3     57
## 23 ComplexTreatmentWell       2      2     37
## 24            Diathermy       2      2     48
## 25                  IFC       2      2     45
## 26              Thermal       2      2     37
## 27                Botox       1      1     31
## 28     ComplexTreatment       1      1     54
## 29      Electromagnetic       1      1     40
## 30              Glucose       1      1     17
## 31            Injection       1      1     24
## 32                 NMES       1      1     20
```

###### Summary Table: Sensitivity Outcome

###### GRADE Table: Sensitivity Outcome

GRADE Summary (Pain Short-Term)

| Intervention | total\_N | Risk of bias | Imprecision | Inconsistency | Indirectness | Publication bias |
| --- | --- | --- | --- | --- | --- | --- |
| Anaesthetic | 255 | Serious risk of bias | No serious imprecision | No serious inconsistency | No serious indirectness | Undetected |
| ComplexTreatmentWell | 311 | No serious risk of bias | No serious imprecision | No serious inconsistency | No serious indirectness | Undetected |
| ComplexTreatmentPoor | 478 | No serious risk of bias | No serious imprecision | No serious inconsistency | No serious indirectness | Undetected |
| LowIntensity | 482 | No serious risk of bias | No serious imprecision | No serious inconsistency | No serious indirectness | Undetected |
| Hyaluroinic | 223 | No serious risk of bias | No serious imprecision | No serious inconsistency | No serious indirectness | Undetected |
| HighIntensity | 756 | No serious risk of bias | No serious imprecision | No serious inconsistency | No serious indirectness | Undetected |

###### Summary Figure: Sensitivity Outcome

**Figure 1. Effect size plots** Shows the
minimal‐intervention benchmark alongside interventions that included
specific modalities as a primary focus (either alone or as one of
several components). The blue band represents the median and 75%
credible interval for the pooled meta-analytic mean. The shaded
distribution illustrates the predicted effect sizes expected from a
future study using a focused intervention. Black lines indicate the 66%
and 95% middle credible intervals of this predictive
distribution.

##### Mid

###### Mapping

**Figure 1. Focus map without adjunct interventions**  
This figure shows the distribution of the intervention modalities
identified as a focus and their
overlaps.  
  
  
**Figure
2. Focus map including adjunct interventions**  
 This figure
shows the distribution of the intervention modalities identified as a
focus, the adjuncts, and their
overlaps.  
  
  
**Table
1. Summary of interventions**  
 This table summarises the
frequency of intervention modalities identified as a focus across
studies, study arms, and the number of participants included.

```
##         Intervention Studies Groups TotalN
## 1      Strengthening      19     35   1119
## 2                MPR      10     14    489
## 3                ROM       9     17    601
## 4          Injection       8     13    429
## 5           Steroids       8      9    295
## 6   ComplexTreatment       7     10    384
## 7            MTLocal       4      5    124
## 8          Shockwave       4      5    218
## 9       Electrolysis       2      2     50
## 10   FormalEducation       1      3    123
## 11          MTSpinal       1      2     50
## 12    Radiofrequency       1      2     54
## 13 Ultrasonophoresis       1      2     44
## 14          Needling       1      1     24
## 15              TENS       1      1     25
## 16            Taping       1      1     26
```

###### Summary Table

###### GRADE Table

GRADE Summary (Pain Mid-Term)

| Intervention | total\_N | Risk of bias | Imprecision | Inconsistency | Indirectness | Publication bias |
| --- | --- | --- | --- | --- | --- | --- |
| Complex | 384 | No serious risk of bias | Serious imprecision | No serious inconsistency | No serious indirectness | Undetected |
| Shockwave | 218 | No serious risk of bias | Serious imprecision | No serious inconsistency | No serious indirectness | Undetected |
| ROM | 601 | No serious risk of bias | Serious imprecision | No serious inconsistency | No serious indirectness | Undetected |
| Strengthening | 1119 | No serious risk of bias | Serious imprecision | No serious inconsistency | No serious indirectness | Undetected |
| Injection | 429 | No serious risk of bias | Serious imprecision | Serious inconsistency | No serious indirectness | Strongly suspected |
| MT-Local | 124 | No serious risk of bias | Serious imprecision | No serious inconsistency | No serious indirectness | Undetected |
| MPR | 489 | No serious risk of bias | Serious imprecision | No serious inconsistency | No serious indirectness | Undetected |
| Steroids | 295 | No serious risk of bias | Serious imprecision | No serious inconsistency | No serious indirectness | Strongly suspected |

###### Summary Figure

**Figure 1. Effect size plots** Shows the
minimal‐intervention benchmark alongside interventions that included
specific modalities as a primary focus (either alone or as one of
several components). The blue band represents the median and 75%
credible interval for the pooled meta-analytic mean. The shaded
distribution illustrates the predicted effect sizes expected from a
future study using a focused intervention. Black lines indicate the 66%
and 95% middle credible intervals of this predictive
distribution.

###### Risk of bias

Risk of Bias (Pain Mid-Term)

| Study | RoB |
| --- | --- |
| Arias-Buría et al. 2017 | LOW |
| Østerås et al. 2010 | LOW |
| Marzetti et al. 2014 | LOW |
| Eliason et al. 2021 | HIGH |
| Avendaño-Coy et al. 2022 | LOW |
| Schydlowsky et al. 2022 | LOW |
| Eliason et al. 2022 | LOW |
| Raeesi et al. 2023 | LOW |
| Dubé et al. 2023 | LOW |
| Dejaco et al. 2017 | HIGH |
| Liu et al. 2024 | LOW |
| Hotta et al. 2020 | LOW |
| Ketola et al. 2009 | LOW |
| Kesikburun et al. 2013 | LOW |
| Góngora-Rodríguez et al. 2024 | LOW |
| Paavola et al. 2018 | LOW |
| Mulligan et al. 2016 | LOW |
| de Miguel Valtierra et al. 2018 | LOW |
| Cavaggion et al. 2024 | LOW |
| de Oliveira et al. 2021 | LOW |
| Engebretsen et al. 2009 | LOW |
| Jo et al. 2020 | LOW |
| Turgut et al. 2025 | LOW |
| Penning et al. 2012 | LOW |
| Cole et al. 2018 | LOW |
| Rossi et al. 2024 | LOW |
| Esmaily et al. 2022 | LOW |
| Ko et al. 2024 | LOW |
| Apivatgaroon et al. 2023 | LOW |
| Daghiani et al. 2023 | LOW |
| Bennell et al. 2010 | LOW |
| Hunter et al. 2022 | LOW |
| Kolk et al. 2013 | LOW |
| Kvalvaag et al. 2017 | SOME |

###### Network Analysis

**Figure 1. Interactive network plot**  
 This figure
shows the pairwise comparisons between single-focus interventions that
were included in the corresponding benchmark analysis. Edge thickness
represents the number of pairwise comparisons contributing to each
connection. The nodes can be manipulated, with zooming and translation
using the mouse.

**Insufficient data and network geometry for
analysis.**

###### Mapping: Sensitivity Outcome

**Figure 1. Focus map without adjunct interventions**  
This figure shows the distribution of the intervention modalities
identified as a focus and their
overlaps.  
  
  
**Figure
2. Focus map including adjunct interventions**  
 This figure
shows the distribution of the intervention modalities identified as a
focus, the adjuncts, and their
overlaps.  
  
  
**Table
1. Summary of interventions**  
 This table summarises the
frequency of intervention modalities identified as a focus across
studies, study arms, and the number of participants included.

```
##            Intervention Studies Groups TotalN
## 1         Strengthening      10     18    549
## 2                   MPR      10     11    380
## 3                   ROM       9     17    601
## 4              Steroids       8      9    295
## 5         HighIntensity       7     10    310
## 6          LowIntensity       4      7    260
## 7               MTLocal       4      5    124
## 8             Shockwave       4      5    218
## 9           Hyaluroinic       3      6    192
## 10                  PRP       3      3     93
## 11         Electrolysis       2      2     50
## 12      FormalEducation       1      3    123
## 13             MTSpinal       1      2     50
## 14                Ozone       1      2     72
## 15       Radiofrequency       1      2     54
## 16    Ultrasonophoresis       1      2     44
## 17          Anaesthetic       1      1     55
## 18     ComplexTreatment       1      1     54
## 19 ComplexTreatmentWell       1      1     22
## 20              Glucose       1      1     17
## 21             Needling       1      1     24
## 22                 TENS       1      1     25
## 23               Taping       1      1     26
```

###### Summary Table: Sensitivity Outcome

###### GRADE Table: Sensitivity Outcome

GRADE Summary (Pain Mid-Term)

| Intervention | total\_N | Risk of bias | Imprecision | Inconsistency | Indirectness | Publication bias |
| --- | --- | --- | --- | --- | --- | --- |
| ComplexTreatmentWell | 119 | No serious risk of bias | Serious imprecision | Serious inconsistency | No serious indirectness | Undetected |
| LowIntensity | 260 | No serious risk of bias | No serious imprecision | No serious inconsistency | No serious indirectness | Undetected |
| Hyaluroinic | 192 | No serious risk of bias | Serious imprecision | Serious inconsistency | No serious indirectness | Undetected |
| ComplexTreatmentPoor | 265 | No serious risk of bias | Serious imprecision | No serious inconsistency | No serious indirectness | Undetected |
| HighIntensity | 310 | No serious risk of bias | Serious imprecision | No serious inconsistency | No serious indirectness | Undetected |

###### Summary Figure: Sensitivity Outcome

**Figure 1. Effect size plots** Shows the
minimal‐intervention benchmark alongside interventions that included
specific modalities as a primary focus (either alone or as one of
several components). The blue band represents the median and 75%
credible interval for the pooled meta-analytic mean. The shaded
distribution illustrates the predicted effect sizes expected from a
future study using a focused intervention. Black lines indicate the 66%
and 95% middle credible intervals of this predictive
distribution.

##### Long

###### Mapping

**Figure 1. Focus map without adjunct interventions**  
This figure shows the distribution of the intervention modalities
identified as a focus and their
overlaps.  
  
  
**Figure
2. Focus map including adjunct interventions**  
 This figure
shows the distribution of the intervention modalities identified as a
focus, the adjuncts, and their
overlaps.  
  
  
**Table
1. Summary of interventions**  
 This table summarises the
frequency of intervention modalities identified as a focus across
studies, study arms, and the number of participants included.

```
##        Intervention Studies Groups TotalN
## 1     Strengthening       9     14    519
## 2         Injection       4      7    234
## 3               MPR       3      4    229
## 4         Shockwave       3      4    178
## 5          Steroids       3      4    181
## 6               ROM       2      4    174
## 7          MTSpinal       2      3     94
## 8           MTLocal       2      2     69
## 9          Needling       2      2     42
## 10 ComplexTreatment       1      2    134
## 11     Electrolysis       1      1     18
```

###### Summary Table

###### GRADE Table

GRADE Summary (Pain Long-Term)

| Intervention | total\_N | Risk of bias | Imprecision | Inconsistency | Indirectness | Publication bias |
| --- | --- | --- | --- | --- | --- | --- |
| ROM | 174 | No serious risk of bias | No serious imprecision | No serious inconsistency | No serious indirectness | Undetected |
| Injection | 234 | No serious risk of bias | No serious imprecision | No serious inconsistency | No serious indirectness | Strongly suspected |
| Strengthening | 519 | No serious risk of bias | Serious imprecision | No serious inconsistency | No serious indirectness | Undetected |
| Steroids | 181 | No serious risk of bias | Serious imprecision | No serious inconsistency | No serious indirectness | Strongly suspected |
| MPR | 229 | No serious risk of bias | Serious imprecision | No serious inconsistency | No serious indirectness | Undetected |
| Shockwave | 178 | No serious risk of bias | Serious imprecision | No serious inconsistency | No serious indirectness | Undetected |

###### Summary Figure

**Figure 1. Effect size plots** Shows the
minimal‐intervention benchmark alongside interventions that included
specific modalities as a primary focus (either alone or as one of
several components). The blue band represents the median and 75%
credible interval for the pooled meta-analytic mean. The shaded
distribution illustrates the predicted effect sizes expected from a
future study using a focused intervention. Black lines indicate the 66%
and 95% middle credible intervals of this predictive
distribution.

###### Risk of bias

Risk of Bias (Pain Long-Term)

| Study | RoB |
| --- | --- |
| Arias-Buría et al. 2017 | LOW |
| Østerås et al. 2010 | LOW |
| Rodríguez-Huguet et al. 2020 | LOW |
| Ketola et al. 2009 | LOW |
| Haahr et al. 2005 | LOW |
| Kesikburun et al. 2013 | LOW |
| Paavola et al. 2018 | LOW |
| Kromer et al. 2014 | LOW |
| Hallgren et al. 2014 | LOW |
| Turgut et al. 2025 | LOW |
| Rossi et al. 2024 | LOW |
| Ko et al. 2024 | LOW |
| Engebretsen et al. 2011 | LOW |
| Kvalvaag et al. 2018 | LOW |

###### Network Analysis

**Figure 1. Interactive network plot**  
 This figure
shows the pairwise comparisons between single-focus interventions that
were included in the corresponding benchmark analysis. Edge thickness
represents the number of pairwise comparisons contributing to each
connection. The nodes can be manipulated, with zooming and translation
using the mouse.

**Insufficient data and network geometry for
analysis.**

###### Mapping: Sensitivity Outcome

**Figure 1. Focus map without adjunct interventions**  
This figure shows the distribution of the intervention modalities
identified as a focus and their
overlaps.  
  
  
**Figure
2. Focus map including adjunct interventions**  
 This figure
shows the distribution of the intervention modalities identified as a
focus, the adjuncts, and their
overlaps.  
  
  
**Table
1. Summary of interventions**  
 This table summarises the
frequency of intervention modalities identified as a focus across
studies, study arms, and the number of participants included.

```
##     Intervention Studies Groups TotalN
## 1  Strengthening       5      7    238
## 2   LowIntensity       3      6    231
## 3      Shockwave       3      4    178
## 4       Steroids       3      4    181
## 5            MPR       3      3    161
## 6            ROM       2      4    174
## 7       MTSpinal       2      3     94
## 8        MTLocal       2      2     69
## 9       Needling       2      2     42
## 10           PRP       2      2     70
## 11   Hyaluroinic       1      3     92
## 12         Ozone       1      2     72
## 13  Electrolysis       1      1     18
## 14 HighIntensity       1      1     50
```

###### Summary Table: Sensitivity Outcome

###### GRADE Table: Sensitivity Outcome

GRADE Summary (Pain Long-Term)

| Intervention | total\_N | Risk of bias | Imprecision | Inconsistency | Indirectness | Publication bias |
| --- | --- | --- | --- | --- | --- | --- |
| LowIntensity | 231 | No serious risk of bias | Serious imprecision | No serious inconsistency | No serious indirectness | Undetected |

###### Summary Figure: Sensitivity Outcome

**Figure 1. Effect size plots** Shows the
minimal‐intervention benchmark alongside interventions that included
specific modalities as a primary focus (either alone or as one of
several components). The blue band represents the median and 75%
credible interval for the pooled meta-analytic mean. The shaded
distribution illustrates the predicted effect sizes expected from a
future study using a focused intervention. Black lines indicate the 66%
and 95% middle credible intervals of this predictive
distribution.

#### Function

##### Short

###### Mapping

**Figure 1. Focus map without adjunct interventions**  
This figure shows the distribution of the intervention modalities
identified as a focus and their
overlaps.  
  
  
**Figure
2. Focus map including adjunct interventions**  
 This figure
shows the distribution of the intervention modalities identified as a
focus, the adjuncts, and their
overlaps.  
  
  
**Table
1. Summary of interventions**  
 This table summarises the
frequency of intervention modalities identified as a focus across
studies, study arms, and the number of participants included.

```
##         Intervention Studies Groups TotalN
## 1      Strengthening      62    102   3253
## 2                ROM      34     56   1798
## 3           Steroids      23     31   1288
## 4   ComplexTreatment      20     28   1122
## 5                MPR      19     25    795
## 6            MTLocal      19     24    649
## 7          Injection      13     19    643
## 8          Shockwave       9     10    337
## 9         Ultrasound       8      9    221
## 10          MTSpinal       7      8    293
## 11            Taping       6      6    158
## 12          Needling       5      7    347
## 13             Laser       5      5    133
## 14   FormalEducation       3      5    299
## 15 Ultrasonophoresis       3      4     84
## 16              TENS       2      4     95
## 17    Radiofrequency       2      3     81
## 18         Diathermy       2      2     34
## 19      Electrolysis       2      2     50
## 20   Electromagnetic       1      1     40
## 21               IFC       1      1     30
## 22              NMES       1      1     20
## 23              TDCS       1      1     20
## 24           Thermal       1      1     23
```

###### Summary Table

###### GRADE Table

GRADE Summary (Function Short-Term)

| Intervention | total\_N | Risk of bias | Imprecision | Inconsistency | Indirectness | Publication bias |
| --- | --- | --- | --- | --- | --- | --- |
| Laser | 133 | No serious risk of bias | No serious imprecision | Serious inconsistency | No serious indirectness | Undetected |
| Taping | 158 | No serious risk of bias | No serious imprecision | Serious inconsistency | No serious indirectness | Undetected |
| Needling | 347 | No serious risk of bias | No serious imprecision | No serious inconsistency | No serious indirectness | Strongly suspected |
| Ultrasound | 221 | No serious risk of bias | No serious imprecision | Serious inconsistency | No serious indirectness | Undetected |
| Complex | 1122 | No serious risk of bias | No serious imprecision | No serious inconsistency | No serious indirectness | Undetected |
| MT-Local | 649 | No serious risk of bias | No serious imprecision | No serious inconsistency | No serious indirectness | Undetected |
| MPR | 795 | No serious risk of bias | No serious imprecision | No serious inconsistency | No serious indirectness | Undetected |
| MT-Spinal | 293 | No serious risk of bias | No serious imprecision | No serious inconsistency | No serious indirectness | Undetected |
| ROM | 1798 | No serious risk of bias | No serious imprecision | No serious inconsistency | No serious indirectness | Undetected |
| Steroids | 1288 | No serious risk of bias | No serious imprecision | No serious inconsistency | No serious indirectness | Strongly suspected |
| Strengthening | 3253 | No serious risk of bias | No serious imprecision | No serious inconsistency | No serious indirectness | Undetected |
| Shockwave | 337 | No serious risk of bias | No serious imprecision | Serious inconsistency | No serious indirectness | Undetected |
| Injection | 643 | No serious risk of bias | No serious imprecision | No serious inconsistency | No serious indirectness | Strongly suspected |
| Education | 299 | No serious risk of bias | Serious imprecision | No serious inconsistency | No serious indirectness | Undetected |

###### Summary Figure

**Figure 1. Effect size plots** Shows the
minimal‐intervention benchmark alongside interventions that included
specific modalities as a primary focus (either alone or as one of
several components). The blue band represents the median and 75%
credible interval for the pooled meta-analytic mean. The shaded
distribution illustrates the predicted effect sizes expected from a
future study using a focused intervention. Black lines indicate the 66%
and 95% middle credible intervals of this predictive
distribution.

###### Risk of bias

Risk of Bias (Function Short-Term)

| Study | RoB |
| --- | --- |
| Boudreau et al. 2019 | LOW |
| Arias-Buría et al. 2017 | LOW |
| Østerås et al. 2010 | LOW |
| Tauqeer et al. 2024 | LOW |
| Struyf et al. 2013 | LOW |
| Marzetti et al. 2014 | LOW |
| Camargo et al. 2015 | LOW |
| Michener et al. 2024 | LOW |
| Ehsani et al. 2024 | LOW |
| Lewis et al. 2017 | LOW |
| Eliason et al. 2021 | LOW |
| Kara et al. 2024 | LOW |
| Blume et al. 2015 | LOW |
| Letafatkar et al. 2021 | LOW |
| Avendaño-Coy et al. 2022 | LOW |
| de Oliveira et al. 2022 | LOW |
| Abu EL Kasem et al. 2024 | HIGH |
| Pekgöz et al. 2020 | LOW |
| Heron et al. 2017 | LOW |
| Schydlowsky et al. 2022 | LOW |
| Dupuis et al. 2018 | LOW |
| Eliason et al. 2022 | LOW |
| Raeesi et al. 2023 | LOW |
| Dubé et al. 2023 | LOW |
| Dejaco et al. 2017 | LOW |
| Clausen et al. 2021 | SOME |
| Kandemir et al. 2024 | HIGH |
| Gutiérrez-Espinoza et al. 2023a | SOME |
| Liu et al. 2024 | LOW |
| Ludewig & Borstad 2003 | LOW |
| Beltrán et al. 2024 | LOW |
| Hotta et al. 2020 | LOW |
| Ketola et al. 2009 | LOW |
| Dilek et al. 2016 | HIGH |
| Haahr et al. 2005 | LOW |
| Elgendy et al. 2023 | LOW |
| Li et al. 2024 | LOW |
| Sen et al. 2023 | LOW |
| Türksan et al. 2024 | LOW |
| San Segundo et al. 2008 | LOW |
| Kesikburun et al. 2013 | LOW |
| Juul-Kristensen et al. 2019 | LOW |
| Lombardi et al. 2008 | LOW |
| Hopewell et al. 2021 | LOW |
| Kamonseki et al. 2023 | LOW |
| Kulakli et al. 2020 | LOW |
| Nguyen et al. 2024 | LOW |
| Gutiérrez Espinoza et al. 2023b | LOW |
| Góngora-Rodríguez et al. 2024 | LOW |
| Johansson et al. 2011 | LOW |
| Mulligan et al. 2016 | HIGH |
| Ingwersen et al. 2017 | LOW |
| de Miguel Valtierra et al. 2018 | LOW |
| Arias-Buriá et al. 2015 | LOW |
| Bal et al. 2009 | LOW |
| Kromer et al. 2014 | LOW |
| Delgado-Gil et al. 2025 | LOW |
| Holmgren et al. 2012 | LOW |
| Buccioli et al. 2025 | SOME |
| Qureshi et al. 2024 | LOW |
| Cavaggion et al. 2024 | HIGH |
| Saleh et al. 2025 | LOW |
| Tahran & Yeşilyaprak 2020 | HIGH |
| Granviken & Vasseljen 2015 | HIGH |
| Szczurko et al. 2009 | LOW |
| Giombini et al. 2006 | HIGH |
| Babaei-Ghazani et al. 2019 | LOW |
| Ogbeivor et al. 2019 | LOW |
| Karthikeyan et al. 2010 | LOW |
| Jo et al. 2020 | LOW |
| Dogu et al. 2012 | LOW |
| Turgut et al. 2025 | LOW |
| Lee et al. 2011 | LOW |
| Akbari et al. 2020 | LOW |
| Apivatgaroon et al. 2023 | LOW |
| Ebadi et al. 2023 | LOW |
| Penning et al. 2012 | LOW |
| Ekeberg et al. 2009 | LOW |
| Daghiani et al. 2023 | LOW |
| Wang et al. 2019 | LOW |
| Cole et al. 2016 | LOW |
| Nazary-Moghadam et al. 2025 | LOW |
| Crawshaw et al. 2010 | SOME |
| Bennell et al. 2010 | SOME |
| Sırlan et al. 2025 | SOME |
| de Oliveira et al. 2021 | SOME |
| Kim et al. 2020 | LOW |
| Engebretsen et al. 2009 | LOW |
| Dunning et al. 2021 | LOW |
| Ishaq et al. 2024 | LOW |
| Baeske et al. 2024 | LOW |
| Hunter et al. 2022 | LOW |
| Khandaloo et al. 2025 | LOW |
| Chou et al. 2010 | LOW |
| Esmaily et al. 2022 | HIGH |
| Ottani et al. 2024 | HIGH |
| Ko et al. 2024 | LOW |
| Kim et al. 2025 | LOW |
| Santamato et al. 2009a | LOW |
| Li et al. 2017 | LOW |
| Kolk et al. 2013 | LOW |
| Kvalvaag et al. 2017 | LOW |
| Johansson et al. 2005 | SOME |
| Yavuz et al. 2014 | LOW |
| Santamato et al. 2009b | SOME |
| Vinuesa-Montoya et al. 2017 | LOW |
| Taik et al. 2022 | LOW |
| Kocyigit et al. 2016 | LOW |
| Shin et al. 2021 | LOW |
| Kibar et al. 2017 | SOME |
| Akbaş et al. 2025 | LOW |

###### Network Analysis

**Figure 1. Interactive network plot**  
 This figure
shows the pairwise comparisons between single-focus interventions that
were included in the corresponding benchmark analysis. Edge thickness
represents the number of pairwise comparisons contributing to each
connection. The nodes can be manipulated, with zooming and translation
using the mouse.

**Table 1. Network analysis results**  
 This table
summarises the mean pairwise difference in Function (SMD) between each
single-focus intervention and the reference treatment, strengthening.
Negative values indicate that strengthening is superior; positive values
indicate that strengthening is inferior.

**Table 2. Network ranking results**  
 This table
summarises the ranking of interventions from the network analysis,
including the mean rank and its uncertainty, the probability of ranking
1st and in the top 3, and the SUCRA (Surface Under the Cumulative
Ranking Curve) value with its uncertainty.  
  

| treatment | mean\_rank | rank\_lower | rank\_upper | mean\_sucra | sucra\_lower | sucra\_upper | prob\_rank\_1 | prob\_rank\_1\_to\_3 |
| --- | --- | --- | --- | --- | --- | --- | --- | --- |
| Laser | 4.00 | 1 | 12 | 0.79 | 0.21 | 1.00 | 0.25 | 0.56 |
| MTLocal | 4.94 | 1 | 12 | 0.72 | 0.21 | 1.00 | 0.12 | 0.39 |
| Complex | 5.03 | 1 | 12 | 0.71 | 0.21 | 1.00 | 0.11 | 0.37 |
| MPR | 5.28 | 1 | 12 | 0.69 | 0.21 | 1.00 | 0.11 | 0.35 |
| Ultrasound | 5.41 | 1 | 13 | 0.69 | 0.14 | 1.00 | 0.08 | 0.34 |
| Needling | 7.23 | 1 | 14 | 0.56 | 0.07 | 1.00 | 0.12 | 0.28 |
| ROM | 7.80 | 1 | 14 | 0.51 | 0.07 | 1.00 | 0.03 | 0.15 |
| Taping | 8.16 | 1 | 14 | 0.49 | 0.07 | 1.00 | 0.07 | 0.20 |
| Steroids | 8.30 | 3 | 13 | 0.48 | 0.14 | 0.86 | 0.00 | 0.04 |
| MTSpinal | 8.32 | 1 | 14 | 0.48 | 0.07 | 1.00 | 0.06 | 0.17 |
| FormalEducation | 9.09 | 2 | 14 | 0.42 | 0.07 | 0.93 | 0.02 | 0.09 |
| Strengthening | 10.08 | 7 | 13 | 0.35 | 0.14 | 0.57 | 0.00 | 0.00 |
| Injection | 10.41 | 4 | 14 | 0.33 | 0.07 | 0.79 | 0.00 | 0.02 |
| Shockwave | 11.13 | 3 | 15 | 0.28 | 0.00 | 0.86 | 0.01 | 0.04 |
| Control | 14.82 | 13 | 15 | 0.01 | 0.00 | 0.14 | 0.00 | 0.00 |

**Table 3. Network GRADE analysis**  
 This table
summarises the GRADE assessment of the network analysis.   
  

GRADE Summary of Findings (Function, Short-Term)

| Analysis | total\_N | Risk of bias | Imprecision | Inconsistency | Indirectness | Publication bias |
| --- | --- | --- | --- | --- | --- | --- |
| Function (short-term) | 2333 | No serious risk of bias | Very serious imprecision | Some concern of inconsistency | No serious indirectness | Unclear |
|  |  | Most contributing studies were randomised trials with acceptable methods; risk of bias was unlikely to materially affect relative treatment comparisons. | Relative treatment effects were small compared with typical within-arm Function reductions, with wide credible intervals crossing no difference. Ranking estimates were unstable, with several interventions spanning most possible rank positions and wide SUCRA intervals. | Between-study heterogeneity was moderate (τ = 0.33; 95% CrI 0.18 to 0.53 on the smd-change scale). Residual deviance modestly exceeded the number of observations (posterior mean residual deviance ≈ 70 vs 60 arm-level observations). However, allowing for inconsistency did not improve model fit (Base model mean Dres = 70.1, Inconsistency model mean Dres = 69.3, ΔDres = 0.8), indicating agreement between direct and indirect evidence. | Included studies directly addressed the population, interventions, and Function outcomes of interest; indirect comparisons were clinically plausible. | Formal assessment of small-study effects was limited by the number of studies contributing to each comparison. |

###### Mapping: Sensitivity Outcome

**Figure 1. Focus map without adjunct interventions**  
This figure shows the distribution of the intervention modalities
identified as a focus and their
overlaps.  
  
  
**Figure
2. Focus map including adjunct interventions**  
 This figure
shows the distribution of the intervention modalities identified as a
focus, the adjuncts, and their
overlaps.  
  
  
**Table
1. Summary of interventions**  
 This table summarises the
frequency of intervention modalities identified as a focus across
studies, study arms, and the number of participants included.

```
##            Intervention Studies Groups TotalN
## 1                   ROM      34     55   1746
## 2         Strengthening      34     51   1671
## 3              Steroids      23     31   1288
## 4         HighIntensity      19     31   1012
## 5               MTLocal      19     24    649
## 6                   MPR      19     21    649
## 7          LowIntensity      11     19    554
## 8             Shockwave       9     10    337
## 9            Ultrasound       8      9    221
## 10             MTSpinal       7      8    293
## 11               Taping       6      6    158
## 12             Needling       5      7    347
## 13                Laser       5      5    133
## 14          Hyaluroinic       4      7    223
## 15      FormalEducation       3      5    299
## 16          Anaesthetic       3      4    203
## 17    Ultrasonophoresis       3      4     84
## 18                 TENS       2      4     95
## 19     ComplexTreatment       2      3    259
## 20                Ozone       2      3     87
## 21       Radiofrequency       2      3     81
## 22 ComplexTreatmentWell       2      2     37
## 23            Diathermy       2      2     34
## 24         Electrolysis       2      2     50
## 25                  PRP       2      2     45
## 26                Botox       1      1     31
## 27      Electromagnetic       1      1     40
## 28                  IFC       1      1     30
## 29            Injection       1      1     24
## 30                 NMES       1      1     20
## 31                 TDCS       1      1     20
## 32            Tenoxicam       1      1     30
## 33              Thermal       1      1     23
```

###### Summary Table: Sensitivity Outcome

###### GRADE Table: Sensitivity Outcome

GRADE Summary (Function Short-Term)

| Intervention | total\_N | Risk of bias | Imprecision | Inconsistency | Indirectness | Publication bias |
| --- | --- | --- | --- | --- | --- | --- |
| ComplexTreatmentWell | 523 | No serious risk of bias | No serious imprecision | No serious inconsistency | No serious indirectness | Undetected |
| ComplexTreatmentPoor | 599 | No serious risk of bias | No serious imprecision | Serious inconsistency | No serious indirectness | Undetected |
| Hyaluroinic | 223 | No serious risk of bias | No serious imprecision | No serious inconsistency | No serious indirectness | Undetected |
| LowIntensity | 554 | No serious risk of bias | No serious imprecision | Serious inconsistency | No serious indirectness | Undetected |
| HighIntensity | 1012 | No serious risk of bias | No serious imprecision | No serious inconsistency | No serious indirectness | Undetected |
| Anaesthetic | 203 | Very serious risk of bias | Serious imprecision | Serious inconsistency | No serious indirectness | Undetected |

###### Summary Figure: Sensitivity Outcome

**Figure 1. Effect size plots** Shows the
minimal‐intervention benchmark alongside interventions that included
specific modalities as a primary focus (either alone or as one of
several components). The blue band represents the median and 75%
credible interval for the pooled meta-analytic mean. The shaded
distribution illustrates the predicted effect sizes expected from a
future study using a focused intervention. Black lines indicate the 66%
and 95% middle credible intervals of this predictive
distribution.

##### Mid

###### Mapping

**Figure 1. Focus map without adjunct interventions**  
This figure shows the distribution of the intervention modalities
identified as a focus and their
overlaps.  
  
  
**Figure
2. Focus map including adjunct interventions**  
 This figure
shows the distribution of the intervention modalities identified as a
focus, the adjuncts, and their
overlaps.  
  
  
**Table
1. Summary of interventions**  
 This table summarises the
frequency of intervention modalities identified as a focus across
studies, study arms, and the number of participants included.

```
##         Intervention Studies Groups TotalN
## 1      Strengthening      25     44   1731
## 2                ROM      12     24    919
## 3                MPR      12     17    560
## 4   ComplexTreatment      11     16    802
## 5           Steroids       9     11    692
## 6          Injection       5      9    313
## 7            MTLocal       5      6    176
## 8          Shockwave       4      5    218
## 9    FormalEducation       2      4    266
## 10          MTSpinal       2      3    102
## 11          Needling       2      3    170
## 12      Electrolysis       2      2     50
## 13    Radiofrequency       1      2     54
## 14 Ultrasonophoresis       1      2     44
## 15              TENS       1      1     25
## 16            Taping       1      1     26
```

###### Summary Table

###### GRADE Table

GRADE Summary (Function Mid-Term)

| Intervention | total\_N | Risk of bias | Imprecision | Inconsistency | Indirectness | Publication bias |
| --- | --- | --- | --- | --- | --- | --- |
| MT-Local | 176 | No serious risk of bias | Serious imprecision | No serious inconsistency | No serious indirectness | Undetected |
| MT-Spinal | 102 | No serious risk of bias | Serious imprecision | No serious inconsistency | No serious indirectness | Undetected |
| Complex | 802 | No serious risk of bias | Serious imprecision | Serious inconsistency | No serious indirectness | Undetected |
| Needling | 170 | No serious risk of bias | Serious imprecision | No serious inconsistency | No serious indirectness | Strongly suspected |
| MPR | 560 | No serious risk of bias | Serious imprecision | Serious inconsistency | No serious indirectness | Undetected |
| ROM | 919 | No serious risk of bias | Serious imprecision | No serious inconsistency | No serious indirectness | Undetected |
| Education | 266 | No serious risk of bias | Serious imprecision | No serious inconsistency | No serious indirectness | Undetected |
| Shockwave | 218 | No serious risk of bias | Serious imprecision | No serious inconsistency | No serious indirectness | Undetected |
| Strengthening | 1731 | No serious risk of bias | Serious imprecision | No serious inconsistency | No serious indirectness | Undetected |
| Steroids | 692 | No serious risk of bias | Serious imprecision | No serious inconsistency | No serious indirectness | Strongly suspected |
| Injection | 313 | No serious risk of bias | Serious imprecision | No serious inconsistency | No serious indirectness | Strongly suspected |

###### Summary Figure

**Figure 1. Effect size plots** Shows the
minimal‐intervention benchmark alongside interventions that included
specific modalities as a primary focus (either alone or as one of
several components). The blue band represents the median and 75%
credible interval for the pooled meta-analytic mean. The shaded
distribution illustrates the predicted effect sizes expected from a
future study using a focused intervention. Black lines indicate the 66%
and 95% middle credible intervals of this predictive
distribution.

###### Risk of bias

Risk of Bias (Function Mid-Term)

| Study | RoB |
| --- | --- |
| Østerås et al. 2010 | LOW |
| Marzetti et al. 2014 | LOW |
| Michener et al. 2024 | LOW |
| Lewis et al. 2017 | LOW |
| Eliason et al. 2021 | LOW |
| Avendaño-Coy et al. 2022 | HIGH |
| Schydlowsky et al. 2022 | LOW |
| Eliason et al. 2022 | LOW |
| Raeesi et al. 2023 | LOW |
| Dubé et al. 2023 | LOW |
| Dejaco et al. 2017 | LOW |
| Clausen et al. 2021 | HIGH |
| Liu et al. 2024 | LOW |
| Hotta et al. 2020 | LOW |
| Ketola et al. 2009 | LOW |
| Haahr et al. 2005 | LOW |
| Kesikburun et al. 2013 | LOW |
| Hopewell et al. 2021 | LOW |
| Góngora-Rodríguez et al. 2024 | LOW |
| Johansson et al. 2011 | LOW |
| Paavola et al. 2018 | LOW |
| Mulligan et al. 2016 | LOW |
| de Miguel Valtierra et al. 2018 | LOW |
| Buccioli et al. 2025 | LOW |
| Cavaggion et al. 2024 | LOW |
| Granviken & Vasseljen 2015 | LOW |
| de Oliveira et al. 2021 | LOW |
| Engebretsen et al. 2009 | LOW |
| Dickens et al. 2005 | LOW |
| Daghiani et al. 2023 | LOW |
| Bennell et al. 2010 | LOW |
| Crawshaw et al. 2010 | LOW |
| Jo et al. 2020 | LOW |
| Turgut et al. 2025 | LOW |
| Apivatgaroon et al. 2023 | LOW |
| Penning et al. 2012 | LOW |
| Ko et al. 2024 | LOW |
| Hunter et al. 2022 | SOME |
| Kolk et al. 2013 | LOW |
| Kvalvaag et al. 2017 | SOME |

###### Network Analysis

**Figure 1. Interactive network plot**  
 This figure
shows the pairwise comparisons between single-focus interventions that
were included in the corresponding benchmark analysis. Edge thickness
represents the number of pairwise comparisons contributing to each
connection. The nodes can be manipulated, with zooming and translation
using the mouse.

**Table 1. Network analysis results**  
 This table
summarises the mean pairwise difference in Function (SMD) between each
single-focus intervention and the reference treatment, strengthening.
Negative values indicate that strengthening is superior; positive values
indicate that strengthening is inferior.

**Table 2. Network ranking results**  
 This table
summarises the ranking of interventions from the network analysis,
including the mean rank and its uncertainty, the probability of ranking
1st and in the top 3, and the SUCRA (Surface Under the Cumulative
Ranking Curve) value with its uncertainty.  
  

| treatment | mean\_rank | rank\_lower | rank\_upper | mean\_sucra | sucra\_lower | sucra\_upper | prob\_rank\_1 | prob\_rank\_1\_to\_3 |
| --- | --- | --- | --- | --- | --- | --- | --- | --- |
| Complex | 1.94 | 1 | 5 | 0.77 | 0.00 | 1.00 | 0.52 | 0.86 |
| Injection | 1.97 | 1 | 4 | 0.76 | 0.25 | 1.00 | 0.37 | 0.91 |
| Strengthening | 3.40 | 1 | 5 | 0.40 | 0.00 | 1.00 | 0.03 | 0.54 |
| FormalEducation | 3.49 | 1 | 5 | 0.38 | 0.00 | 1.00 | 0.08 | 0.47 |
| Steroids | 4.20 | 2 | 5 | 0.20 | 0.00 | 0.75 | 0.00 | 0.21 |

**Table 3. Network GRADE analysis**  
 This table
summarises the GRADE assessment of the network analysis.   
  

GRADE Summary of Findings (Function, Mid-Term)

| Analysis | total\_N | Risk of bias | Imprecision | Inconsistency | Indirectness | Publication bias |
| --- | --- | --- | --- | --- | --- | --- |
| Function (Mid-term) | 1159 | No serious risk of bias | Very serious imprecision | No serious inconsistency | No serious indirectness | Unclear |
|  |  | Most contributing studies were randomised trials with acceptable methods; risk of bias was unlikely to materially affect relative treatment comparisons. | Relative treatment effects were small compared with typical within-arm Function reductions, with wide credible intervals crossing no difference. Ranking estimates were unstable, with several interventions spanning most possible rank positions and wide SUCRA intervals. | Between-study heterogeneity was small (τ = 0.10; 95% CrI 0.01 to 0.29 on the smd-change scale). Global model fit was adequate (posterior mean residual deviance ≈ 15 vs 17 arm-level observations). Allowing for inconsistency did not improve model fit (Base model mean Dres = 15.3, Inconsistency model mean Dres = 15.1, ΔDres = 0.2), indicating agreement between direct and indirect evidence. | Included studies directly addressed the population, interventions, and Function outcomes of interest; indirect comparisons were clinically plausible. | Formal assessment of small-study effects was limited by the number of studies contributing to each comparison. |

###### Mapping: Sensitivity Outcome

```
##            Intervention Studies Groups TotalN
## 1         Strengthening      13     22    917
## 2                   ROM      12     23    867
## 3                   MPR      12     14    451
## 4         HighIntensity       9     13    461
## 5              Steroids       9     11    692
## 6          LowIntensity       5      9    353
## 7               MTLocal       5      6    176
## 8             Shockwave       4      5    218
## 9      ComplexTreatment       3      4    292
## 10      FormalEducation       2      4    266
## 11          Hyaluroinic       2      4    143
## 12             MTSpinal       2      3    102
## 13             Needling       2      3    170
## 14         Electrolysis       2      2     50
## 15                  PRP       2      2     43
## 16                Ozone       1      2     72
## 17       Radiofrequency       1      2     54
## 18    Ultrasonophoresis       1      2     44
## 19          Anaesthetic       1      1     55
## 20 ComplexTreatmentWell       1      1     22
## 21                 TENS       1      1     25
## 22               Taping       1      1     26
```

###### Summary Table: Sensitivity Outcome

###### GRADE Table: Sensitivity Outcome

GRADE Summary (Function Mid-Term)

| Intervention | total\_N | Risk of bias | Imprecision | Inconsistency | Indirectness | Publication bias |
| --- | --- | --- | --- | --- | --- | --- |
| ComplexTreatmentPoor | 438 | No serious risk of bias | Serious imprecision | Serious inconsistency | No serious indirectness | Undetected |
| LowIntensity | 353 | No serious risk of bias | Serious imprecision | Serious inconsistency | No serious indirectness | Undetected |
| ComplexTreatmentWell | 364 | No serious risk of bias | Serious imprecision | No serious inconsistency | No serious indirectness | Undetected |
| HighIntensity | 461 | Serious risk of bias | Serious imprecision | No serious inconsistency | No serious indirectness | Undetected |
| Hyaluroinic | 143 | No serious risk of bias | Serious imprecision | Serious inconsistency | No serious indirectness | Undetected |

###### Summary Figure: Sensitivity Outcome

**Figure 1. Effect size plots** Shows the
minimal‐intervention benchmark alongside interventions that included
specific modalities as a primary focus (either alone or as one of
several components). The blue band represents the median and 75%
credible interval for the pooled meta-analytic mean. The shaded
distribution illustrates the predicted effect sizes expected from a
future study using a focused intervention. Black lines indicate the 66%
and 95% middle credible intervals of this predictive
distribution.

##### Long

###### Mapping

**Figure 1. Focus map without adjunct interventions**  
This figure shows the distribution of the intervention modalities
identified as a focus and their
overlaps.  
  
  
**Figure
2. Focus map including adjunct interventions**  
 This figure
shows the distribution of the intervention modalities identified as a
focus, the adjuncts, and their
overlaps.  
  
  
**Table
1. Summary of interventions**  
 This table summarises the
frequency of intervention modalities identified as a focus across
studies, study arms, and the number of participants included.

```
##        Intervention Studies Groups TotalN
## 1     Strengthening      11     17    892
## 2               ROM       4      9    432
## 3          Steroids       4      6    501
## 4         Injection       3      6    184
## 5  ComplexTreatment       3      5    304
## 6               MPR       3      4    229
## 7          MTSpinal       3      4    146
## 8         Shockwave       3      4    178
## 9           MTLocal       3      3    121
## 10         Needling       2      3    160
## 11  FormalEducation       1      1    143
```

###### Summary Table

###### GRADE Table

GRADE Summary (Function Long-Term)

| Intervention | total\_N | Risk of bias | Imprecision | Inconsistency | Indirectness | Publication bias |
| --- | --- | --- | --- | --- | --- | --- |
| Needling | 160 | No serious risk of bias | Serious imprecision | No serious inconsistency | No serious indirectness | Strongly suspected |
| Strengthening | 892 | No serious risk of bias | Serious imprecision | Serious inconsistency | No serious indirectness | Undetected |
| MT-Spinal | 146 | No serious risk of bias | Serious imprecision | No serious inconsistency | No serious indirectness | Undetected |
| ROM | 432 | No serious risk of bias | Serious imprecision | Serious inconsistency | No serious indirectness | Undetected |
| Complex | 304 | No serious risk of bias | Serious imprecision | Serious inconsistency | No serious indirectness | Undetected |
| MT-Local | 121 | Serious risk of bias | Serious imprecision | Serious inconsistency | No serious indirectness | Undetected |
| MPR | 229 | No serious risk of bias | Serious imprecision | Serious inconsistency | No serious indirectness | Undetected |
| Steroids | 501 | No serious risk of bias | Serious imprecision | No serious inconsistency | No serious indirectness | Strongly suspected |
| Injection | 184 | No serious risk of bias | Serious imprecision | Serious inconsistency | No serious indirectness | Strongly suspected |
| Shockwave | 178 | No serious risk of bias | Serious imprecision | Serious inconsistency | No serious indirectness | Undetected |

###### Summary Figure

**Figure 1. Effect size plots** Shows the
minimal‐intervention benchmark alongside interventions that included
specific modalities as a primary focus (either alone or as one of
several components). The blue band represents the median and 75%
credible interval for the pooled meta-analytic mean. The shaded
distribution illustrates the predicted effect sizes expected from a
future study using a focused intervention. Black lines indicate the 66%
and 95% middle credible intervals of this predictive
distribution.

###### Risk of bias

Risk of Bias (Function Long-Term)

| Study | RoB |
| --- | --- |
| Østerås et al. 2010 | LOW |
| Michener et al. 2024 | LOW |
| Lewis et al. 2017 | LOW |
| Ketola et al. 2009 | LOW |
| Haahr et al. 2005 | LOW |
| Kesikburun et al. 2013 | LOW |
| Hopewell et al. 2021 | LOW |
| Johansson et al. 2011 | LOW |
| Paavola et al. 2018 | LOW |
| Kromer et al. 2014 | LOW |
| Hallgren et al. 2014 | LOW |
| Turgut et al. 2025 | LOW |
| Ko et al. 2024 | LOW |
| Engebretsen et al. 2011 | LOW |
| Hunter et al. 2022 | LOW |
| Kvalvaag et al. 2018 | LOW |

###### Network Analysis

**Figure 1. Interactive network plot**  
 This figure
shows the pairwise comparisons between single-focus interventions that
were included in the corresponding benchmark analysis. Edge thickness
represents the number of pairwise comparisons contributing to each
connection. The nodes can be manipulated, with zooming and translation
using the mouse.


visNetwork

**Table 1. Network analysis results**  
 This table
summarises the mean pairwise difference in Function (SMD) between each
single-focus intervention and the reference treatment, strengthening.
Negative values indicate that strengthening is superior; positive values
indicate that strengthening is inferior.

**Table 2. Network ranking results**  
 This table
summarises the ranking of interventions from the network analysis,
including the mean rank and its uncertainty, the probability of ranking
1st and in the top 3, and the SUCRA (Surface Under the Cumulative
Ranking Curve) value with its uncertainty.  
  

| treatment | mean\_rank | rank\_lower | rank\_upper | mean\_sucra | sucra\_lower | sucra\_upper | prob\_rank\_1 | prob\_rank\_1\_to\_3 |
| --- | --- | --- | --- | --- | --- | --- | --- | --- |
| Complex | 2.45 | 1 | 5 | 0.64 | 0 | 1 | 0.39 | 0.72 |
| Injection | 2.46 | 1 | 5 | 0.64 | 0 | 1 | 0.34 | 0.74 |
| Strengthening | 2.89 | 1 | 5 | 0.53 | 0 | 1 | 0.09 | 0.71 |
| FormalEducation | 3.41 | 1 | 5 | 0.40 | 0 | 1 | 0.15 | 0.47 |
| Steroids | 3.79 | 1 | 5 | 0.30 | 0 | 1 | 0.03 | 0.35 |

**Table 3. Network GRADE analysis**  
 This table
summarises the GRADE assessment of the network analysis.   
  

GRADE Summary of Findings (Function, Long-Term)

| Analysis | total\_N | Risk of bias | Imprecision | Inconsistency | Indirectness | Publication bias |
| --- | --- | --- | --- | --- | --- | --- |
| Function (Long-term) | 890 | No serious risk of bias | Very serious imprecision | No serious inconsistency | No serious indirectness | Unclear |
|  |  | Most contributing studies were randomised trials with acceptable methods; risk of bias was unlikely to materially affect relative treatment comparisons. | Relative treatment effects were small compared with typical within-arm Function reductions, with wide credible intervals crossing no difference. Ranking estimates were unstable, with several interventions spanning most possible rank positions and wide SUCRA intervals. | Between-study heterogeneity was moderate (τ = 0.29; 95% CrI 0.06 to 0.64 on the smd-change scale). Global model fit was adequate (posterior mean residual deviance ≈ 10 vs 10 arm-level observations). Allowing for inconsistency did not improve model fit (Base model mean Dres = 10.4, Inconsistency model mean Dres = 9.9, ΔDres = 0.5), indicating agreement between direct and indirect evidence. | Included studies directly addressed the population, interventions, and Function outcomes of interest; indirect comparisons were clinically plausible. | Formal assessment of small-study effects was limited by the number of studies contributing to each comparison. |

###### Mapping: Sensitivity Outcome

**Figure 1. Focus map without adjunct interventions**  
This figure shows the distribution of the intervention modalities
identified as a focus and their
overlaps.  
  
  
**Figure
2. Focus map including adjunct interventions**  
 This figure
shows the distribution of the intervention modalities identified as a
focus, the adjuncts, and their
overlaps.  
  
  
**Table
1. Summary of interventions**  
 This table summarises the
frequency of intervention modalities identified as a focus across
studies, study arms, and the number of participants included.

```
##       Intervention Studies Groups TotalN
## 1    Strengthening       8     11    604
## 2              ROM       4      8    380
## 3         Steroids       4      6    501
## 4     LowIntensity       3      6    288
## 5         MTSpinal       3      4    146
## 6        Shockwave       3      4    178
## 7              MPR       3      3    161
## 8          MTLocal       3      3    121
## 9         Needling       2      3    160
## 10     Hyaluroinic       1      3     92
## 11           Ozone       1      2     72
## 12 FormalEducation       1      1    143
## 13   HighIntensity       1      1     50
## 14             PRP       1      1     20
```

###### Summary Table: Sensitivity Outcome

###### GRADE Table: Sensitivity Outcome

GRADE Summary (Function Long-Term)

| Intervention | total\_N | Risk of bias | Imprecision | Inconsistency | Indirectness | Publication bias |
| --- | --- | --- | --- | --- | --- | --- |
| ComplexTreatmentPoor | 252 | No serious risk of bias | Serious imprecision | Serious inconsistency | No serious indirectness | Undetected |
| LowIntensity | 288 | No serious risk of bias | Serious imprecision | Serious inconsistency | No serious indirectness | Undetected |

###### Summary Figure: Sensitivity Outcome

**Figure 1. Effect size plots** Shows the
minimal‐intervention benchmark alongside interventions that included
specific modalities as a primary focus (either alone or as one of
several components). The blue band represents the median and 75%
credible interval for the pooled meta-analytic mean. The shaded
distribution illustrates the predicted effect sizes expected from a
future study using a focused intervention. Black lines indicate the 66%
and 95% middle credible intervals of this predictive
distribution.

#### QOL

##### Short

###### Mapping

**Figure 1. Focus map without adjunct interventions**  
This figure shows the distribution of the intervention modalities
identified as a focus and their
overlaps.  
  
  
**Figure
2. Focus map including adjunct interventions**  
 This figure
shows the distribution of the intervention modalities identified as a
focus, the adjuncts, and their
overlaps.  
  
  
**Table
1. Summary of interventions**  
 This table summarises the
frequency of intervention modalities identified as a focus across
studies, study arms, and the number of participants included.

```
##         Intervention Studies Groups TotalN
## 1      Strengthening      17     24    862
## 2           Steroids       8     11    655
## 3                ROM       7     10    287
## 4          Injection       5      8    277
## 5   ComplexTreatment       5      5    160
## 6                MPR       4      4    141
## 7  Ultrasonophoresis       3      4     84
## 8             Taping       3      3     57
## 9    FormalEducation       2      4    255
## 10    Radiofrequency       2      3     81
## 11           MTLocal       2      2     36
## 12         Shockwave       2      2     36
## 13         Diathermy       1      1     20
## 14   Electromagnetic       1      1     40
## 15              NMES       1      1     20
## 16          Needling       1      1     42
## 17              TDCS       1      1     20
## 18           Thermal       1      1     23
```

###### Summary Table

###### GRADE Table

GRADE Summary (QOL Short-Term)

| Intervention | total\_N | Risk of bias | Imprecision | Inconsistency | Indirectness | Publication bias |
| --- | --- | --- | --- | --- | --- | --- |
| MPR | 141 | No serious risk of bias | No serious imprecision | Serious inconsistency | No serious indirectness | Undetected |
| Injection | 277 | No serious risk of bias | No serious imprecision | No serious inconsistency | No serious indirectness | Strongly suspected |
| Complex | 160 | No serious risk of bias | Serious imprecision | Serious inconsistency | No serious indirectness | Undetected |
| ROM | 287 | No serious risk of bias | No serious imprecision | No serious inconsistency | No serious indirectness | Undetected |
| Education | 255 | No serious risk of bias | Serious imprecision | Serious inconsistency | No serious indirectness | Undetected |
| Steroids | 655 | No serious risk of bias | No serious imprecision | No serious inconsistency | No serious indirectness | Strongly suspected |
| Strengthening | 862 | No serious risk of bias | No serious imprecision | No serious inconsistency | No serious indirectness | Undetected |

###### Summary Figure

**Figure 1. Effect size plots** Shows the
minimal‐intervention benchmark alongside interventions that included
specific modalities as a primary focus (either alone or as one of
several components). The blue band represents the median and 75%
credible interval for the pooled meta-analytic mean. The shaded
distribution illustrates the predicted effect sizes expected from a
future study using a focused intervention. Black lines indicate the 66%
and 95% middle credible intervals of this predictive
distribution.

###### Risk of bias

Risk of Bias (QOL Short-Term)

| Study | RoB |
| --- | --- |
| Boudreau et al. 2019 | LOW |
| Avendaño-Coy et al. 2022 | LOW |
| Pekgöz et al. 2020 | LOW |
| Dupuis et al. 2018 | SOME |
| Dubé et al. 2023 | LOW |
| Raeesi et al. 2023 | LOW |
| Clausen et al. 2021 | LOW |
| Kandemir et al. 2024 | LOW |
| Liu et al. 2024 | LOW |
| Dilek et al. 2016 | LOW |
| Kesikburun et al. 2013 | LOW |
| Lombardi et al. 2008 | LOW |
| Hopewell et al. 2021 | LOW |
| Johansson et al. 2011 | LOW |
| Holmgren et al. 2012 | LOW |
| Qureshi et al. 2024 | LOW |
| Cavaggion et al. 2024 | LOW |
| Turgut et al. 2025 | LOW |
| Apivatgaroon et al. 2023 | LOW |
| Ekeberg et al. 2009 | LOW |
| Daghiani et al. 2023 | SOME |
| Nazary-Moghadam et al. 2025 | HIGH |
| Szczurko et al. 2009 | LOW |
| Esmaily et al. 2022 | LOW |
| Kim et al. 2025 | LOW |
| Bennell et al. 2010 | LOW |
| de Oliveira et al. 2021 | LOW |

###### Network Analysis

**Figure 1. Interactive network plot**  
 This figure
shows the pairwise comparisons between single-focus interventions that
were included in the corresponding benchmark analysis. Edge thickness
represents the number of pairwise comparisons contributing to each
connection. The nodes can be manipulated, with zooming and translation
using the mouse.

**Table 1. Network analysis results**  
 This table
summarises the mean pairwise difference in QOL (SMD) between each
single-focus intervention and the reference treatment, strengthening.
Negative values indicate that strengthening is superior; positive values
indicate that strengthening is inferior.

**Table 2. Network ranking results**  
 This table
summarises the ranking of interventions from the network analysis,
including the mean rank and its uncertainty, the probability of ranking
1st and in the top 3, and the SUCRA (Surface Under the Cumulative
Ranking Curve) value with its uncertainty.  
  

| treatment | mean\_rank | rank\_lower | rank\_upper | mean\_sucra | sucra\_lower | sucra\_upper | prob\_rank\_1 | prob\_rank\_1\_to\_3 |
| --- | --- | --- | --- | --- | --- | --- | --- | --- |
| Complex | 1.81 | 1 | 5 | 0.84 | 0.2 | 1.0 | 0.59 | 0.89 |
| Steroids | 3.46 | 1 | 6 | 0.51 | 0.0 | 1.0 | 0.09 | 0.53 |
| FormalEducation | 3.72 | 1 | 6 | 0.46 | 0.0 | 1.0 | 0.13 | 0.46 |
| Control | 3.87 | 1 | 6 | 0.43 | 0.0 | 1.0 | 0.06 | 0.43 |
| Injection | 4.05 | 1 | 6 | 0.39 | 0.0 | 1.0 | 0.11 | 0.39 |
| Strengthening | 4.09 | 2 | 6 | 0.38 | 0.0 | 0.8 | 0.01 | 0.29 |

**Table 3. Network GRADE analysis**  
 This table
summarises the GRADE assessment of the network analysis.   
  

GRADE Summary of Findings (QOL, Short-Term)

| Analysis | total\_N | Risk of bias | Imprecision | Inconsistency | Indirectness | Publication bias |
| --- | --- | --- | --- | --- | --- | --- |
| QOL (Short-term) | 921 | No serious risk of bias | Very serious imprecision | Some concern of inconsistency | No serious indirectness | Unclear |
|  |  | Most contributing studies were randomised trials with acceptable methods; risk of bias was unlikely to materially affect relative treatment comparisons. | Relative treatment effects were small compared with typical within-arm QOL reductions, with wide credible intervals crossing no difference. Ranking estimates were unstable, with several interventions spanning most possible rank positions and wide SUCRA intervals. | Between-study heterogeneity was moderate to substantial (τ = 0.45; 95% CrI 0.16 to 0.86 on the smd-change scale). Residual deviance modestly exceeded the number of observations (posterior mean residual deviance ≈ 16 vs 13 arm-level observations). However, allowing for inconsistency did not improve model fit (Base model mean Dres = 15.7, Inconsistency model mean Dres = 14.9, ΔDres = 0.8), indicating agreement between direct and indirect evidence. | Included studies directly addressed the population, interventions, and QOL outcomes of interest; indirect comparisons were clinically plausible. | Formal assessment of small-study effects was limited by the number of studies contributing to each comparison. |

###### Mapping: Sensitivity Outcome

**Figure 1. Focus map without adjunct interventions**  
This figure shows the distribution of the intervention modalities
identified as a focus and their
overlaps.  
  
  
**Figure
2. Focus map including adjunct interventions**  
 This figure
shows the distribution of the intervention modalities identified as a
focus, the adjuncts, and their
overlaps.  
  
  
**Table
1. Summary of interventions**  
 This table summarises the
frequency of intervention modalities identified as a focus across
studies, study arms, and the number of participants included.

```
##            Intervention Studies Groups TotalN
## 1         Strengthening       8     13    461
## 2              Steroids       8     11    655
## 3                   ROM       7     10    287
## 4         HighIntensity       7      8    327
## 5                   MPR       4      4    141
## 6     Ultrasonophoresis       3      4     84
## 7          LowIntensity       3      3     74
## 8                Taping       3      3     57
## 9       FormalEducation       2      4    255
## 10       Radiofrequency       2      3     81
## 11              MTLocal       2      2     36
## 12            Shockwave       2      2     36
## 13          Anaesthetic       1      2    106
## 14          Hyaluroinic       1      2     55
## 15                Ozone       1      2     72
## 16     ComplexTreatment       1      1     54
## 17 ComplexTreatmentWell       1      1     22
## 18            Diathermy       1      1     20
## 19      Electromagnetic       1      1     40
## 20            Injection       1      1     24
## 21                 NMES       1      1     20
## 22             Needling       1      1     42
## 23                  PRP       1      1     20
## 24                 TDCS       1      1     20
## 25              Thermal       1      1     23
```

###### Summary Table: Sensitivity Outcome

###### GRADE Table: Sensitivity Outcome

GRADE Summary (QOL Short-Term)

| Intervention | total\_N | Risk of bias | Imprecision | Inconsistency | Indirectness | Publication bias |
| --- | --- | --- | --- | --- | --- | --- |
| HighIntensity | 327 | No serious risk of bias | No serious imprecision | No serious inconsistency | No serious indirectness | Undetected |

###### Summary Figure: Sensitivity Outcome

**Figure 1. Effect size plots** Shows the
minimal‐intervention benchmark alongside interventions that included
specific modalities as a primary focus (either alone or as one of
several components). The blue band represents the median and 75%
credible interval for the pooled meta-analytic mean. The shaded
distribution illustrates the predicted effect sizes expected from a
future study using a focused intervention. Black lines indicate the 66%
and 95% middle credible intervals of this predictive
distribution.

##### Mid

###### Mapping

**Figure 1. Focus map without adjunct interventions**  
This figure shows the distribution of the intervention modalities
identified as a focus and their
overlaps.  
  
  
**Figure
2. Focus map including adjunct interventions**  
 This figure
shows the distribution of the intervention modalities identified as a
focus, the adjuncts, and their
overlaps.  
  
  
**Table
1. Summary of interventions**  
 This table summarises the
frequency of intervention modalities identified as a focus across
studies, study arms, and the number of participants included.

```
##         Intervention Studies Groups TotalN
## 1      Strengthening       9     13    564
## 2           Steroids       5      7    478
## 3   ComplexTreatment       4      4    150
## 4                MPR       4      4    141
## 5                ROM       3      4    114
## 6    FormalEducation       2      4    257
## 7          Injection       2      3     92
## 8     Radiofrequency       1      2     54
## 9  Ultrasonophoresis       1      2     44
## 10          Needling       1      1     42
## 11            Taping       1      1     26
```

###### Summary Table

###### GRADE Table

GRADE Summary (QOL Mid-Term)

| Intervention | total\_N | Risk of bias | Imprecision | Inconsistency | Indirectness | Publication bias |
| --- | --- | --- | --- | --- | --- | --- |
| ROM | 114 | No serious risk of bias | No serious imprecision | Serious inconsistency | No serious indirectness | Undetected |
| MPR | 141 | No serious risk of bias | No serious imprecision | Serious inconsistency | No serious indirectness | Undetected |
| Education | 257 | No serious risk of bias | Serious imprecision | Serious inconsistency | No serious indirectness | Undetected |
| Complex | 150 | No serious risk of bias | Serious imprecision | Serious inconsistency | No serious indirectness | Undetected |
| Strengthening | 564 | No serious risk of bias | No serious imprecision | No serious inconsistency | No serious indirectness | Undetected |
| Steroids | 478 | No serious risk of bias | Serious imprecision | No serious inconsistency | No serious indirectness | Strongly suspected |

###### Summary Figure

**Figure 1. Effect size plots** Shows the
minimal‐intervention benchmark alongside interventions that included
specific modalities as a primary focus (either alone or as one of
several components). The blue band represents the median and 75%
credible interval for the pooled meta-analytic mean. The shaded
distribution illustrates the predicted effect sizes expected from a
future study using a focused intervention. Black lines indicate the 66%
and 95% middle credible intervals of this predictive
distribution.

###### Risk of bias

Risk of Bias (QOL Mid-Term)

| Study | RoB |
| --- | --- |
| Avendaño-Coy et al. 2022 | LOW |
| Dubé et al. 2023 | LOW |
| Raeesi et al. 2023 | LOW |
| Clausen et al. 2021 | LOW |
| Liu et al. 2024 | LOW |
| Kesikburun et al. 2013 | LOW |
| Hopewell et al. 2021 | LOW |
| Johansson et al. 2011 | LOW |
| Cavaggion et al. 2024 | LOW |
| Turgut et al. 2025 | LOW |
| Apivatgaroon et al. 2023 | LOW |
| Daghiani et al. 2023 | LOW |
| Bennell et al. 2010 | LOW |
| de Oliveira et al. 2021 | SOME |

###### Network Analysis

**Figure 1. Interactive network plot**  
 This figure
shows the pairwise comparisons between single-focus interventions that
were included in the corresponding benchmark analysis. Edge thickness
represents the number of pairwise comparisons contributing to each
connection. The nodes can be manipulated, with zooming and translation
using the mouse.

**Insufficient data and network geometry for
analysis.**

###### Mapping: Sensitivity Outcome

**Figure 1. Focus map without adjunct interventions**  
This figure shows the distribution of the intervention modalities
identified as a focus and their
overlaps.  
  
  
**Figure
2. Focus map including adjunct interventions**  
 This figure
shows the distribution of the intervention modalities identified as a
focus, the adjuncts, and their
overlaps.  
  
  
**Table
1. Summary of interventions**  
 This table summarises the
frequency of intervention modalities identified as a focus across
studies, study arms, and the number of participants included.

```
##            Intervention Studies Groups TotalN
## 1              Steroids       5      7    478
## 2         Strengthening       4      7    306
## 3         HighIntensity       4      4    214
## 4                   MPR       4      4    141
## 5                   ROM       3      4    114
## 6       FormalEducation       2      4    257
## 7          LowIntensity       2      2     44
## 8                 Ozone       1      2     72
## 9        Radiofrequency       1      2     54
## 10    Ultrasonophoresis       1      2     44
## 11     ComplexTreatment       1      1     54
## 12 ComplexTreatmentWell       1      1     22
## 13             Needling       1      1     42
## 14                  PRP       1      1     20
## 15               Taping       1      1     26
```

###### Summary Table: Sensitivity Outcome

###### GRADE Table: Sensitivity Outcome

GRADE Summary (QOL Mid-Term)

| Intervention | total\_N | Risk of bias | Imprecision | Inconsistency | Indirectness | Publication bias |
| --- | --- | --- | --- | --- | --- | --- |
| HighIntensity | 214 | No serious risk of bias | Serious imprecision | Serious inconsistency | No serious indirectness | Undetected |

###### Summary Figure: Sensitivity Outcome

**Figure 1. Effect size plots** Shows the
minimal‐intervention benchmark alongside interventions that included
specific modalities as a primary focus (either alone or as one of
several components). The blue band represents the median and 75%
credible interval for the pooled meta-analytic mean. The shaded
distribution illustrates the predicted effect sizes expected from a
future study using a focused intervention. Black lines indicate the 66%
and 95% middle credible intervals of this predictive
distribution.

#### S17. Active-intervention study characteristics

Embedded PDF not supported.
Download
S17 PDF

#### S18. Bayesian predictive meta-analyses comparing active interventions with minimal-intervention benchmarking (Sensitvity 1)

### Outcomes

#### Pain

##### Short

###### Mapping

**Figure 1. Focus map without adjunct interventions**  
This figure shows the distribution of the intervention modalities
identified as a focus and their
overlaps.  
  
  
**Figure
2. Focus map including adjunct interventions**  
 This figure
shows the distribution of the intervention modalities identified as a
focus, the adjuncts, and their
overlaps.  
  
  
**Table
1. Summary of interventions**  
 This table summarises the
frequency of intervention modalities identified as a single focus across
studies, study arms, and the number of participants included.

```
##         Intervention Studies Groups TotalN
## 18     Strengthening      17     24    638
## 17          Steroids      14     20    587
## 7          Injection      11     13    422
## 10           MTLocal       8      9    254
## 8              Laser       5      5    133
## 9                MPR       5      5    149
## 16         Shockwave       5      5    165
## 23        Ultrasound       5      5     97
## 1   ComplexTreatment       3      3     91
## 15               ROM       3      3     75
## 19            Taping       3      3     72
## 22 Ultrasonophoresis       2      3     57
## 2          Diathermy       2      2     48
## 21           Thermal       2      2     37
## 12          Needling       1      2     54
## 3       Electrolysis       1      1     17
## 5    FormalEducation       1      1     41
## 6                IFC       1      1     30
## 11          MTSpinal       1      1     25
## 14    Radiofrequency       1      1     27
## 4    Electromagnetic       0      0      0
## 13              NMES       0      0      0
## 20              TENS       0      0      0
```

###### Summary Table

###### GRADE Table

| Intervention | total\_N | Risk of bias | Imprecision | Inconsistency | Indirectness | Publication bias |
| --- | --- | --- | --- | --- | --- | --- |
| Laser | 133 | No serious risk of bias | No serious imprecision | No serious inconsistency | No serious indirectness | Undetected |
| Shockwave | 165 | No serious risk of bias | No serious imprecision | Serious inconsistency | No serious indirectness | Undetected |
| MPR | 149 | No serious risk of bias | No serious imprecision | No serious inconsistency | No serious indirectness | Undetected |
| Injection | 422 | No serious risk of bias | No serious imprecision | Serious inconsistency | No serious indirectness | Strongly suspected |
| MTLocal | 254 | No serious risk of bias | No serious imprecision | No serious inconsistency | No serious indirectness | Undetected |
| Strengthening | 638 | No serious risk of bias | No serious imprecision | No serious inconsistency | No serious indirectness | Undetected |
| Steroids | 587 | No serious risk of bias | No serious imprecision | No serious inconsistency | No serious indirectness | Strongly suspected |

###### Summary Figure

**Figure 1. Effect size plots** Shows the
minimal‐intervention benchmark alongside interventions that included
specific modalities as a single focus. The blue band represents the
median and 75% credible interval for the pooled meta-analytic mean. The
shaded distribution illustrates the predicted effect sizes expected from
a future study using a focused intervention. Black lines indicate the
66% and 95% middle credible intervals of this predictive
distribution.

##### Mid

###### Mapping

**Figure 1. Focus map without adjunct interventions**  
This figure shows the distribution of the intervention modalities
identified as a focus and their
overlaps.  
  
  
**Figure
2. Focus map including adjunct interventions**  
 This figure
shows the distribution of the intervention modalities identified as a
focus, the adjuncts, and their
overlaps.  
  
  
**Table
1. Summary of interventions**  
 This table summarises the
frequency of intervention modalities identified as a single focus across
studies, study arms, and the number of participants included.

```
##         Intervention Studies Groups TotalN
## 4          Injection       7      9    292
## 12          Steroids       7      8    274
## 13     Strengthening       5      7    225
## 1   ComplexTreatment       2      2     76
## 5                MPR       2      2     78
## 11         Shockwave       2      2     87
## 16 Ultrasonophoresis       1      2     44
## 3    FormalEducation       1      1     41
## 7           MTSpinal       1      1     25
## 14            Taping       1      1     26
## 2       Electrolysis       0      0      0
## 6            MTLocal       0      0      0
## 8           Needling       0      0      0
## 9     Radiofrequency       0      0      0
## 10               ROM       0      0      0
## 15              TENS       0      0      0
```

###### Summary Table

###### GRADE Table

| Intervention | total\_N | Risk of bias | Imprecision | Inconsistency | Indirectness | Publication bias |
| --- | --- | --- | --- | --- | --- | --- |
| Strengthening | 225 | No serious risk of bias | Serious imprecision | No serious inconsistency | No serious indirectness | Undetected |
| Injection | 292 | No serious risk of bias | Serious imprecision | Serious inconsistency | No serious indirectness | Strongly suspected |
| Steroids | 274 | No serious risk of bias | Serious imprecision | No serious inconsistency | No serious indirectness | Strongly suspected |

###### Summary Figure

**Figure 1. Effect size plots** Shows the
minimal‐intervention benchmark alongside interventions that included
specific modalities as a single focus. The blue band represents the
median and 75% credible interval for the pooled meta-analytic mean. The
shaded distribution illustrates the predicted effect sizes expected from
a future study using a focused intervention. Black lines indicate the
66% and 95% middle credible intervals of this predictive
distribution.

##### Long

###### Mapping

**Figure 1. Focus map without adjunct interventions**  
This figure shows the distribution of the intervention modalities
identified as a focus and their
overlaps.  
  
  
**Figure
2. Focus map including adjunct interventions**  
 This figure
shows the distribution of the intervention modalities identified as a
focus, the adjuncts, and their
overlaps.  
  
  
**Table
1. Summary of interventions**  
 This table summarises the
frequency of intervention modalities identified as a single focus across
studies, study arms, and the number of participants included.

```
##        Intervention Studies Groups TotalN
## 3         Injection       3      4    152
## 11    Strengthening       3      4    149
## 10         Steroids       2      2     86
## 4               MPR       1      1     52
## 6          MTSpinal       1      1     25
## 9         Shockwave       1      1     52
## 1  ComplexTreatment       0      0      0
## 2      Electrolysis       0      0      0
## 5           MTLocal       0      0      0
## 7          Needling       0      0      0
## 8               ROM       0      0      0
```

###### Summary Table

###### GRADE Table

| Intervention | total\_N | Risk of bias | Imprecision | Inconsistency | Indirectness | Publication bias |
| --- | --- | --- | --- | --- | --- | --- |
| Injection | 152 | No serious risk of bias | Serious imprecision | Serious inconsistency | No serious indirectness | Strongly suspected |
| Strengthening | 149 | No serious risk of bias | Serious imprecision | Serious inconsistency | No serious indirectness | Undetected |

###### Summary Figure

**Figure 1. Effect size plots** Shows the
minimal‐intervention benchmark alongside interventions that included
specific modalities as a single focus. The blue band represents the
median and 75% credible interval for the pooled meta-analytic mean. The
shaded distribution illustrates the predicted effect sizes expected from
a future study using a focused intervention. Black lines indicate the
66% and 95% middle credible intervals of this predictive
distribution.

#### Function

##### Short

###### Mapping

**Figure 1. Focus map without adjunct interventions**  
This figure shows the distribution of the intervention modalities
identified as a focus and their
overlaps.  
  
  
**Figure
2. Focus map including adjunct interventions**  
 This figure
shows the distribution of the intervention modalities identified as a
focus, the adjuncts, and their
overlaps.  
  
  
**Table
1. Summary of interventions**  
 This table summarises the
frequency of intervention modalities identified as a single focus across
studies, study arms, and the number of participants included.

```
##         Intervention Studies Groups TotalN
## 18     Strengthening      20     30    894
## 17          Steroids      15     22    925
## 7          Injection      10     12    385
## 10           MTLocal       7      9    251
## 24        Ultrasound       6      6    131
## 8              Laser       5      5    133
## 9                MPR       5      5    149
## 1   ComplexTreatment       4      4    197
## 15               ROM       4      4    115
## 16         Shockwave       4      4    149
## 23 Ultrasonophoresis       3      4     84
## 19            Taping       3      3     72
## 12          Needling       2      3     96
## 2          Diathermy       2      2     34
## 5    FormalEducation       2      2    190
## 6                IFC       1      1     30
## 11          MTSpinal       1      1     25
## 14    Radiofrequency       1      1     27
## 20              TDCS       1      1     20
## 22           Thermal       1      1     23
## 3       Electrolysis       0      0      0
## 4    Electromagnetic       0      0      0
## 13              NMES       0      0      0
## 21              TENS       0      0      0
```

###### Summary Table

###### GRADE Table

| Intervention | total\_N | Risk of bias | Imprecision | Inconsistency | Indirectness | Publication bias |
| --- | --- | --- | --- | --- | --- | --- |
| Laser | 133 | No serious risk of bias | No serious imprecision | Serious inconsistency | No serious indirectness | Undetected |
| MTLocal | 251 | No serious risk of bias | No serious imprecision | No serious inconsistency | No serious indirectness | Undetected |
| MPR | 149 | No serious risk of bias | No serious imprecision | Serious inconsistency | No serious indirectness | Undetected |
| Ultrasound | 131 | No serious risk of bias | No serious imprecision | Serious inconsistency | No serious indirectness | Undetected |
| Shockwave | 149 | No serious risk of bias | Serious imprecision | Serious inconsistency | No serious indirectness | Undetected |
| Steroids | 925 | No serious risk of bias | No serious imprecision | No serious inconsistency | No serious indirectness | Strongly suspected |
| Injection | 385 | No serious risk of bias | No serious imprecision | No serious inconsistency | No serious indirectness | Strongly suspected |
| Strengthening | 894 | No serious risk of bias | No serious imprecision | No serious inconsistency | No serious indirectness | Undetected |
| FormalEducation | 190 | No serious risk of bias | Serious imprecision | Serious inconsistency | No serious indirectness | Undetected |
| ROM | 115 | No serious risk of bias | Serious imprecision | No serious inconsistency | No serious indirectness | Undetected |

###### Summary Figure

**Figure 1. Effect size plots** Shows the
minimal‐intervention benchmark alongside interventions that included
specific modalities as a single focus. The blue band represents the
median and 75% credible interval for the pooled meta-analytic mean. The
shaded distribution illustrates the predicted effect sizes expected from
a future study using a focused intervention. Black lines indicate the
66% and 95% middle credible intervals of this predictive
distribution.

##### Mid

###### Mapping

**Figure 1. Focus map without adjunct interventions**  
This figure shows the distribution of the intervention modalities
identified as a focus and their
overlaps.  
  
  
**Figure
2. Focus map including adjunct interventions**  
 This figure
shows the distribution of the intervention modalities identified as a
focus, the adjuncts, and their
overlaps.  
  
  
**Table
1. Summary of interventions**  
 This table summarises the
frequency of intervention modalities identified as a single focus across
studies, study arms, and the number of participants included.

```
##         Intervention Studies Groups TotalN
## 12          Steroids       7      9    575
## 13     Strengthening       6      8    379
## 4          Injection       4      5    176
## 1   ComplexTreatment       4      4    218
## 3    FormalEducation       2      2    184
## 5                MPR       2      2     78
## 11         Shockwave       2      2     87
## 16 Ultrasonophoresis       1      2     44
## 7           MTSpinal       1      1     25
## 14            Taping       1      1     26
## 2       Electrolysis       0      0      0
## 6            MTLocal       0      0      0
## 8           Needling       0      0      0
## 9     Radiofrequency       0      0      0
## 10               ROM       0      0      0
## 15              TENS       0      0      0
```

###### Summary Table

###### GRADE Table

| Intervention | total\_N | Risk of bias | Imprecision | Inconsistency | Indirectness | Publication bias |
| --- | --- | --- | --- | --- | --- | --- |
| Strengthening | 379 | No serious risk of bias | Serious imprecision | No serious inconsistency | No serious indirectness | Undetected |
| FormalEducation | 184 | No serious risk of bias | Serious imprecision | Serious inconsistency | No serious indirectness | Undetected |
| Steroids | 575 | No serious risk of bias | Serious imprecision | No serious inconsistency | No serious indirectness | Strongly suspected |
| Injection | 176 | No serious risk of bias | Serious imprecision | No serious inconsistency | No serious indirectness | Strongly suspected |

###### Summary Figure

**Figure 1. Effect size plots** Shows the
minimal‐intervention benchmark alongside interventions that included
specific modalities as a single focus. The blue band represents the
median and 75% credible interval for the pooled meta-analytic mean. The
shaded distribution illustrates the predicted effect sizes expected from
a future study using a focused intervention. Black lines indicate the
66% and 95% middle credible intervals of this predictive
distribution.

##### Long

###### Mapping

**Figure 1. Focus map without adjunct interventions**  
This figure shows the distribution of the intervention modalities
identified as a focus and their
overlaps.  
  
  
**Figure
2. Focus map including adjunct interventions**  
 This figure
shows the distribution of the intervention modalities identified as a
focus, the adjuncts, and their
overlaps.  
  
  
**Table
1. Summary of interventions**  
 This table summarises the
frequency of intervention modalities identified as a single focus across
studies, study arms, and the number of participants included.

```
##        Intervention Studies Groups TotalN
## 10         Steroids       3      4    406
## 11    Strengthening       3      4    282
## 3         Injection       2      3    102
## 2   FormalEducation       1      1    143
## 4               MPR       1      1     52
## 6          MTSpinal       1      1     25
## 9         Shockwave       1      1     52
## 1  ComplexTreatment       0      0      0
## 5           MTLocal       0      0      0
## 7          Needling       0      0      0
## 8               ROM       0      0      0
```

###### Summary Table

###### GRADE Table

| Intervention | total\_N | Risk of bias | Imprecision | Inconsistency | Indirectness | Publication bias |
| --- | --- | --- | --- | --- | --- | --- |
| Strengthening | 282 | No serious risk of bias | Serious imprecision | Serious inconsistency | No serious indirectness | Undetected |
| Steroids | 406 | No serious risk of bias | Serious imprecision | Serious inconsistency | No serious indirectness | Strongly suspected |
| Injection | 102 | No serious risk of bias | Serious imprecision | Serious inconsistency | No serious indirectness | Strongly suspected |

###### Summary Figure

**Figure 1. Effect size plots** Shows the
minimal‐intervention benchmark alongside interventions that included
specific modalities as a single focus. The blue band represents the
median and 75% credible interval for the pooled meta-analytic mean. The
shaded distribution illustrates the predicted effect sizes expected from
a future study using a focused intervention. Black lines indicate the
66% and 95% middle credible intervals of this predictive
distribution.

#### QOL

##### Short

###### Mapping

**Figure 1. Focus map without adjunct interventions**  
This figure shows the distribution of the intervention modalities
identified as a focus and their
overlaps.  
  
  
**Figure
2. Focus map including adjunct interventions**  
 This figure
shows the distribution of the intervention modalities identified as a
focus, the adjuncts, and their
overlaps.  
  
  
**Table
1. Summary of interventions**  
 This table summarises the
frequency of intervention modalities identified as a single focus across
studies, study arms, and the number of participants included.

```
##         Intervention Studies Groups TotalN
## 13          Steroids       6      8    503
## 14     Strengthening       6      8    333
## 5          Injection       3      5    151
## 18 Ultrasonophoresis       3      4     84
## 1   ComplexTreatment       2      2     76
## 4    FormalEducation       2      2    173
## 15            Taping       2      2     47
## 2          Diathermy       1      1     20
## 6                MPR       1      1     26
## 7            MTLocal       1      1     16
## 10    Radiofrequency       1      1     27
## 11               ROM       1      1     42
## 12         Shockwave       1      1     16
## 16              TDCS       1      1     20
## 17           Thermal       1      1     23
## 3    Electromagnetic       0      0      0
## 8           Needling       0      0      0
## 9               NMES       0      0      0
```

###### Summary Table

###### GRADE Table

| Intervention | total\_N | Risk of bias | Imprecision | Inconsistency | Indirectness | Publication bias |
| --- | --- | --- | --- | --- | --- | --- |
| Injection | 151 | No serious risk of bias | No serious imprecision | No serious inconsistency | No serious indirectness | Strongly suspected |
| Steroids | 503 | No serious risk of bias | No serious imprecision | No serious inconsistency | No serious indirectness | Strongly suspected |
| FormalEducation | 173 | No serious risk of bias | Serious imprecision | Serious inconsistency | No serious indirectness | Undetected |
| Strengthening | 333 | No serious risk of bias | Serious imprecision | No serious inconsistency | No serious indirectness | Undetected |

###### Summary Figure

**Figure 1. Effect size plots** Shows the
minimal‐intervention benchmark alongside interventions that included
specific modalities as a single focus. The blue band represents the
median and 75% credible interval for the pooled meta-analytic mean. The
shaded distribution illustrates the predicted effect sizes expected from
a future study using a focused intervention. Black lines indicate the
66% and 95% middle credible intervals of this predictive
distribution.

##### Mid

###### Mapping

**Figure 1. Focus map without adjunct interventions**  
This figure shows the distribution of the intervention modalities
identified as a focus and their
overlaps.  
  
  
**Figure
2. Focus map including adjunct interventions**  
 This figure
shows the distribution of the intervention modalities identified as a
focus, the adjuncts, and their
overlaps.  
  
  
**Table
1. Summary of interventions**  
 This table summarises the
frequency of intervention modalities identified as a single focus across
studies, study arms, and the number of participants included.

```
##         Intervention Studies Groups TotalN
## 8           Steroids       5      7    478
## 9      Strengthening       2      3    186
## 1   ComplexTreatment       2      2     76
## 2    FormalEducation       2      2    175
## 3          Injection       1      2     72
## 11 Ultrasonophoresis       1      2     44
## 4                MPR       1      1     26
## 10            Taping       1      1     26
## 5           Needling       0      0      0
## 6     Radiofrequency       0      0      0
## 7                ROM       0      0      0
```

###### Summary Table

###### GRADE Table

| Intervention | total\_N | Risk of bias | Imprecision | Inconsistency | Indirectness | Publication bias |
| --- | --- | --- | --- | --- | --- | --- |
| Strengthening | 186 | Serious risk of bias | Serious imprecision | Serious inconsistency | No serious indirectness | Undetected |
| FormalEducation | 175 | No serious risk of bias | Serious imprecision | Serious inconsistency | No serious indirectness | Undetected |
| Steroids | 478 | No serious risk of bias | Serious imprecision | No serious inconsistency | No serious indirectness | Strongly suspected |

###### Summary Figure

**Figure 1. Effect size plots** Shows the
minimal‐intervention benchmark alongside interventions that included
specific modalities as a single focus. The blue band represents the
median and 75% credible interval for the pooled meta-analytic mean. The
shaded distribution illustrates the predicted effect sizes expected from
a future study using a focused intervention. Black lines indicate the
66% and 95% middle credible intervals of this predictive
distribution.

#### S19. Bayesian predictive meta-analyses comparing active interventions with minimal-intervention benchmarking (Sensitvity 2)

### Outcomes

#### Pain

##### Short

###### Mapping

**Figure 1. Focus map without adjunct interventions**  
This figure shows the distribution of the intervention modalities
identified as a focus and their
overlaps.  
  
  
**Figure
2. Focus map including adjunct interventions**  
 This figure
shows the distribution of the intervention modalities identified as a
focus, the adjuncts, and their
overlaps.  
  
  
**Table
1. Summary of interventions**  
 This table summarises the
frequency of intervention modalities identified as a single focus and no
adjuncts across studies, study arms, and the number of participants
included.

```
##         Intervention Studies Groups TotalN
## 18     Strengthening       9     11    254
## 7          Injection       3      4    130
## 1   ComplexTreatment       3      3     91
## 10           MTLocal       3      3     57
## 16         Shockwave       3      3     97
## 15               ROM       2      2     64
## 23        Ultrasound       2      2     47
## 12          Needling       1      2     54
## 3       Electrolysis       1      1     17
## 5    FormalEducation       1      1     41
## 8              Laser       1      1     35
## 9                MPR       1      1     13
## 11          MTSpinal       1      1     25
## 19            Taping       1      1     25
## 21           Thermal       1      1     14
## 2          Diathermy       0      0      0
## 4    Electromagnetic       0      0      0
## 6                IFC       0      0      0
## 13              NMES       0      0      0
## 14    Radiofrequency       0      0      0
## 17          Steroids       0      0      0
## 20              TENS       0      0      0
## 22 Ultrasonophoresis       0      0      0
```

###### Summary Table

###### GRADE Table

| Intervention | total\_N | Risk of bias | Imprecision | Inconsistency | Indirectness | Publication bias |
| --- | --- | --- | --- | --- | --- | --- |
| Injection | 130 | No serious risk of bias | No serious imprecision | No serious inconsistency | No serious indirectness | Strongly suspected |
| Strengthening | 254 | No serious risk of bias | No serious imprecision | No serious inconsistency | No serious indirectness | Undetected |

###### Summary Figure

**Figure 1. Effect size plots** Shows the
minimal‐intervention benchmark alongside interventions that included
specific modalities as a single focus. The blue band represents the
median and 75% credible interval for the pooled meta-analytic mean. The
shaded distribution illustrates the predicted effect sizes expected from
a future study using a focused intervention. Black lines indicate the
66% and 95% middle credible intervals of this predictive
distribution.

##### Mid

###### Mapping

**Figure 1. Focus map without adjunct interventions**  
This figure shows the distribution of the intervention modalities
identified as a focus and their
overlaps.  
  
  
**Figure
2. Focus map including adjunct interventions**  
 This figure
shows the distribution of the intervention modalities identified as a
focus, the adjuncts, and their
overlaps.  
  
  
**Table
1. Summary of interventions**  
 This table summarises the
frequency of intervention modalities identified as a single focus and no
adjuncts across studies, study arms, and the number of participants
included.

```
##         Intervention Studies Groups TotalN
## 4          Injection       2      3     99
## 1   ComplexTreatment       2      2     76
## 13     Strengthening       2      2     53
## 3    FormalEducation       1      1     41
## 7           MTSpinal       1      1     25
## 11         Shockwave       1      1     35
## 2       Electrolysis       0      0      0
## 5                MPR       0      0      0
## 6            MTLocal       0      0      0
## 8           Needling       0      0      0
## 9     Radiofrequency       0      0      0
## 10               ROM       0      0      0
## 12          Steroids       0      0      0
## 14            Taping       0      0      0
## 15              TENS       0      0      0
## 16 Ultrasonophoresis       0      0      0
```

##### Long

###### Mapping

**Figure 1. Focus map without adjunct interventions**  
This figure shows the distribution of the intervention modalities
identified as a focus and their
overlaps.  
  
  
**Figure
2. Focus map including adjunct interventions**  
 This figure
shows the distribution of the intervention modalities identified as a
focus, the adjuncts, and their
overlaps.  
  
  
**Table
1. Summary of interventions**  
 This table summarises the
frequency of intervention modalities identified as a single focus and no
adjuncts across studies, study arms, and the number of participants
included.

```
##        Intervention Studies Groups TotalN
## 3         Injection       1      1     50
## 6          MTSpinal       1      1     25
## 9         Shockwave       1      1     52
## 11    Strengthening       1      1     23
## 1  ComplexTreatment       0      0      0
## 2      Electrolysis       0      0      0
## 4               MPR       0      0      0
## 5           MTLocal       0      0      0
## 7          Needling       0      0      0
## 8               ROM       0      0      0
## 10         Steroids       0      0      0
```

#### Function

##### Short

###### Mapping

**Figure 1. Focus map without adjunct interventions**  
This figure shows the distribution of the intervention modalities
identified as a focus and their
overlaps.  
  
  
**Figure
2. Focus map including adjunct interventions**  
 This figure
shows the distribution of the intervention modalities identified as a
focus, the adjuncts, and their
overlaps.  
  
  
**Table
1. Summary of interventions**  
 This table summarises the
frequency of intervention modalities identified as a single focus and no
adjuncts across studies, study arms, and the number of participants
included.

```
##         Intervention Studies Groups TotalN
## 18     Strengthening      10     12    416
## 1   ComplexTreatment       3      3     91
## 16         Shockwave       3      3     97
## 7          Injection       2      3     80
## 5    FormalEducation       2      2    190
## 10           MTLocal       2      2     36
## 15               ROM       2      2     64
## 24        Ultrasound       2      2     47
## 12          Needling       1      2     54
## 8              Laser       1      1     35
## 9                MPR       1      1     13
## 11          MTSpinal       1      1     25
## 19            Taping       1      1     25
## 2          Diathermy       0      0      0
## 3       Electrolysis       0      0      0
## 4    Electromagnetic       0      0      0
## 6                IFC       0      0      0
## 13              NMES       0      0      0
## 14    Radiofrequency       0      0      0
## 17          Steroids       0      0      0
## 20              TDCS       0      0      0
## 21              TENS       0      0      0
## 22           Thermal       0      0      0
## 23 Ultrasonophoresis       0      0      0
```

###### Summary Table

###### GRADE Table

| Intervention | total\_N | Risk of bias | Imprecision | Inconsistency | Indirectness | Publication bias |
| --- | --- | --- | --- | --- | --- | --- |
| Strengthening | 416 | No serious risk of bias | No serious imprecision | No serious inconsistency | No serious indirectness | Undetected |
| FormalEducation | 190 | No serious risk of bias | Serious imprecision | Serious inconsistency | No serious indirectness | Undetected |

###### Summary Figure

**Figure 1. Effect size plots** Shows the
minimal‐intervention benchmark alongside interventions that included
specific modalities as a single focus. The blue band represents the
median and 75% credible interval for the pooled meta-analytic mean. The
shaded distribution illustrates the predicted effect sizes expected from
a future study using a focused intervention. Black lines indicate the
66% and 95% middle credible intervals of this predictive
distribution.

##### Mid

###### Mapping

**Figure 1. Focus map without adjunct interventions**  
This figure shows the distribution of the intervention modalities
identified as a focus and their
overlaps.  
  
  
**Figure
2. Focus map including adjunct interventions**  
 This figure
shows the distribution of the intervention modalities identified as a
focus, the adjuncts, and their
overlaps.  
  
  
**Table
1. Summary of interventions**  
 This table summarises the
frequency of intervention modalities identified as a single focus and no
adjuncts across studies, study arms, and the number of participants
included.

```
##         Intervention Studies Groups TotalN
## 1   ComplexTreatment       3      3    121
## 13     Strengthening       3      3    207
## 3    FormalEducation       2      2    184
## 7           MTSpinal       1      1     25
## 11         Shockwave       1      1     35
## 2       Electrolysis       0      0      0
## 4          Injection       0      0      0
## 5                MPR       0      0      0
## 6            MTLocal       0      0      0
## 8           Needling       0      0      0
## 9     Radiofrequency       0      0      0
## 10               ROM       0      0      0
## 12          Steroids       0      0      0
## 14            Taping       0      0      0
## 15              TENS       0      0      0
## 16 Ultrasonophoresis       0      0      0
```

###### Summary Table

###### GRADE Table

| Intervention | total\_N | Risk of bias | Imprecision | Inconsistency | Indirectness | Publication bias |
| --- | --- | --- | --- | --- | --- | --- |
| FormalEducation | 184 | No serious risk of bias | Serious imprecision | Serious inconsistency | No serious indirectness | Undetected |
| Strengthening | 207 | No serious risk of bias | Serious imprecision | Serious inconsistency | No serious indirectness | Undetected |

###### Summary Figure

**Figure 1. Effect size plots** Shows the
minimal‐intervention benchmark alongside interventions that included
specific modalities as a single focus. The blue band represents the
median and 75% credible interval for the pooled meta-analytic mean. The
shaded distribution illustrates the predicted effect sizes expected from
a future study using a focused intervention. Black lines indicate the
66% and 95% middle credible intervals of this predictive
distribution.

##### Long

###### Mapping

**Figure 1. Focus map without adjunct interventions**  
This figure shows the distribution of the intervention modalities
identified as a focus and their
overlaps.  
  
  
**Figure
2. Focus map including adjunct interventions**  
 This figure
shows the distribution of the intervention modalities identified as a
focus, the adjuncts, and their
overlaps.  
  
  
**Table
1. Summary of interventions**  
 This table summarises the
frequency of intervention modalities identified as a single focus and no
adjuncts across studies, study arms, and the number of participants
included.

```
##        Intervention Studies Groups TotalN
## 2   FormalEducation       1      1    143
## 6          MTSpinal       1      1     25
## 9         Shockwave       1      1     52
## 11    Strengthening       1      1    153
## 1  ComplexTreatment       0      0      0
## 3         Injection       0      0      0
## 4               MPR       0      0      0
## 5           MTLocal       0      0      0
## 7          Needling       0      0      0
## 8               ROM       0      0      0
## 10         Steroids       0      0      0
```

#### QOL

##### Short

###### Mapping

**Figure 1. Focus map without adjunct interventions**  
This figure shows the distribution of the intervention modalities
identified as a focus and their
overlaps.  
  
  
**Figure
2. Focus map including adjunct interventions**  
 This figure
shows the distribution of the intervention modalities identified as a
focus, the adjuncts, and their
overlaps.  
  
  
**Table
1. Summary of interventions**  
 This table summarises the
frequency of intervention modalities identified as a single focus and no
adjuncts across studies, study arms, and the number of participants
included.

```
##         Intervention Studies Groups TotalN
## 1   ComplexTreatment       2      2     76
## 4    FormalEducation       2      2    173
## 14     Strengthening       2      2    177
## 5          Injection       1      2     55
## 11               ROM       1      1     42
## 2          Diathermy       0      0      0
## 3    Electromagnetic       0      0      0
## 6                MPR       0      0      0
## 7            MTLocal       0      0      0
## 8           Needling       0      0      0
## 9               NMES       0      0      0
## 10    Radiofrequency       0      0      0
## 12         Shockwave       0      0      0
## 13          Steroids       0      0      0
## 15            Taping       0      0      0
## 16              TDCS       0      0      0
## 17           Thermal       0      0      0
## 18 Ultrasonophoresis       0      0      0
```

###### Summary Table

###### GRADE Table

| Intervention | total\_N | Risk of bias | Imprecision | Inconsistency | Indirectness | Publication bias |
| --- | --- | --- | --- | --- | --- | --- |
| FormalEducation | 173 | No serious risk of bias | Serious imprecision | Serious inconsistency | No serious indirectness | Undetected |
| Strengthening | 177 | No serious risk of bias | Serious imprecision | No serious inconsistency | No serious indirectness | Undetected |

###### Summary Figure

**Figure 1. Effect size plots** Shows the
minimal‐intervention benchmark alongside interventions that included
specific modalities as a single focus. The blue band represents the
median and 75% credible interval for the pooled meta-analytic mean. The
shaded distribution illustrates the predicted effect sizes expected from
a future study using a focused intervention. Black lines indicate the
66% and 95% middle credible intervals of this predictive
distribution.

##### Mid

###### Mapping

**Figure 1. Focus map without adjunct interventions**  
This figure shows the distribution of the intervention modalities
identified as a focus and their
overlaps.  
  
  
**Figure
2. Focus map including adjunct interventions**  
 This figure
shows the distribution of the intervention modalities identified as a
focus, the adjuncts, and their
overlaps.  
  
  
**Table
1. Summary of interventions**  
 This table summarises the
frequency of intervention modalities identified as a single focus and no
adjuncts across studies, study arms, and the number of participants
included.

```
##         Intervention Studies Groups TotalN
## 1   ComplexTreatment       2      2     76
## 2    FormalEducation       2      2    175
## 9      Strengthening       1      1    143
## 3          Injection       0      0      0
## 4                MPR       0      0      0
## 5           Needling       0      0      0
## 6     Radiofrequency       0      0      0
## 7                ROM       0      0      0
## 8           Steroids       0      0      0
## 10            Taping       0      0      0
## 11 Ultrasonophoresis       0      0      0
```

###### Summary Table

###### GRADE Table

| Intervention | total\_N | Risk of bias | Imprecision | Inconsistency | Indirectness | Publication bias |
| --- | --- | --- | --- | --- | --- | --- |
| FormalEducation | 175 | No serious risk of bias | Serious imprecision | Serious inconsistency | No serious indirectness | Undetected |

###### Summary Figure

**Figure 1. Effect size plots** Shows the
minimal‐intervention benchmark alongside interventions that included
specific modalities as a single focus. The blue band represents the
median and 75% credible interval for the pooled meta-analytic mean. The
shaded distribution illustrates the predicted effect sizes expected from
a future study using a focused intervention. Black lines indicate the
66% and 95% middle credible intervals of this predictive
distribution.
